# Genetic architecture of cardiac dynamic flow volumes

**DOI:** 10.1101/2022.10.05.22280733

**Authors:** Bruna Gomes, Aditya Singh, Jack W O’Sullivan, David Amar, Mykhailo Kostur, Francois Haddad, Michael Salerno, Victoria N. Parikh, Benjamin Meder, Euan A. Ashley

## Abstract

Cardiac blood flow is a critical determinant of human health. However, definition of its genetic architecture is limited by the technical challenge of capturing dynamic flow volumes from cardiac imaging at scale. We present DeepFlow, a deep learning system to extract cardiac flow and volumes from phase contrast cardiac magnetic resonance imaging. A mixed linear model applied to 37,967 individuals from the UK Biobank reveals novel genome-wide significant associations across cardiac dynamic flow volumes including aortic forward velocity, total left ventricular stroke volume, forward left ventricular flow and aortic regurgitation fraction. Mendelian randomization using CAUSE reveals a causal role for aortic root size in aortic valve regurgitation. The most significant contributing variants (near *ELN, FBN1* and *ULK4)* are implicated in connective tissue and blood pressure pathways. DeepFlow cardiac flow phenotyping at scale, combined with population-level genotyping data in the UK Biobank, reinforces the contribution of connective tissue genes, blood pressure and root size to aortic valve function in the general population.

## Introduction

Cardiac function is central to health and disease. Valvular regurgitation (a loss of integrity of one-way valves between cardiac chambers and vessels) can lead to heart failure, which as a whole, is responsible for 18.6 million annual deaths worldwide^1^. Current therapies for valvular regurgitation are limited largely to surgical or procedural intervention, the risk of which is only warranted once severe regurgitation has led to a decrement in cardiac function. Understanding the genetic underpinnings of cardiac flow volumes is critical to identifying intervenable therapeutic targets for valvular regurgitation prior to this severe stage of disease.

Until recently, the genetic architecture of valvular integrity remained elusive because of technical challenges of phenotyping: noninvasive measurement of cardiac function by echocardiography and cardiac magnetic resonance imaging (MRI) requires labor-intensive manual measurement of end diastolic and end systolic cardiac volumes from select points in the cardiac cycle in order to infer flow from two dimensional representations. This results in noisy phenotyping that makes genetic association studies difficult^2–6^.

Direct measurement of aortic flow by MRI provides an opportunity to assess blood flow while accounting for aortic insufficiency. However, its assessment at scale is also technically challenging: precise segmentation remains a barrier^7^, and data must be obtained from velocity-encoding MRI (phase contrast MRI sequences) that have sequence-specific artifacts. Deep learning, and specifically the adoption of auto-encoders, has enabled a major advance in image segmentation^8^, powering precise extraction of cardiac phenotypes at scale^4^ and enabling the discovery of underlying biology through genetic association ^2,3,9^. No accurate, fully automated, open-source solution for applying these methods to the problem of MRI flow quantification is available.

Here, we combine deep learning strategies with available cardiac MRI and genotyping data from the UK Biobank^10^ to address three major goals: first, we develop an automated software solution to extract cardiac flow and volume from phase contrast magnetic resonance imaging (DeepFlow: **Deep** learning-based aortic blood **Flow** quantification); second, we perform the first genome wide association study of cardiac dynamic flow volumes; finally, we test plausible causal relationships among key anatomic parameters using genetic instruments.

## Results

### DeepFlow reliably extracts cardiac flow metrics at scale

We define cardiac dynamic flow volumes as any moving blood quantity (mL) involving the heart measured over a certain time period (e.g. cardiac cycle, systole, diastole). This includes flow from a cardiac chamber to another cardiac chamber, from a cardiac chamber to a great vessel (e.g. aorta), and from a great vessel back to a cardiac chamber (valvular regurgitation). We concentrated our analysis on several left-sided traits (**Figure 1**) including: total left ventricular stroke volume (the total blood volume that is pumped out the left ventricle during systole); forward left ventricular stroke volume (the blood volume that is pumped from the left ventricle to the aorta during systole); net left ventricular stroke volume (forward left ventricular stroke volume minus aortic valve regurgitant volume); aortic valve regurgitant volume (the blood volume returning from the aorta to the left ventricle in diastole); aortic valve regurgitant fraction (aortic valve regurgitant volume indexed to forward left ventricular stroke volume) and mitral valve regurgitation volume (the blood volume that flows back to the left atrium during systole due to an incompetent mitral valve) by subtracting the forward left ventricular stroke volume from the total left ventricular stroke volume^11^. Additionally, we considered the forward and retrograde peak velocities (cm/s) at the aortic annulus (corresponding to the peak forward (systolic) [Ao_F_Vmax] and peak regurgitant (diastolic) [Ao_R_Vmax] velocities at the aortic annulus, respectively) for analysis.

**Figure 1.**
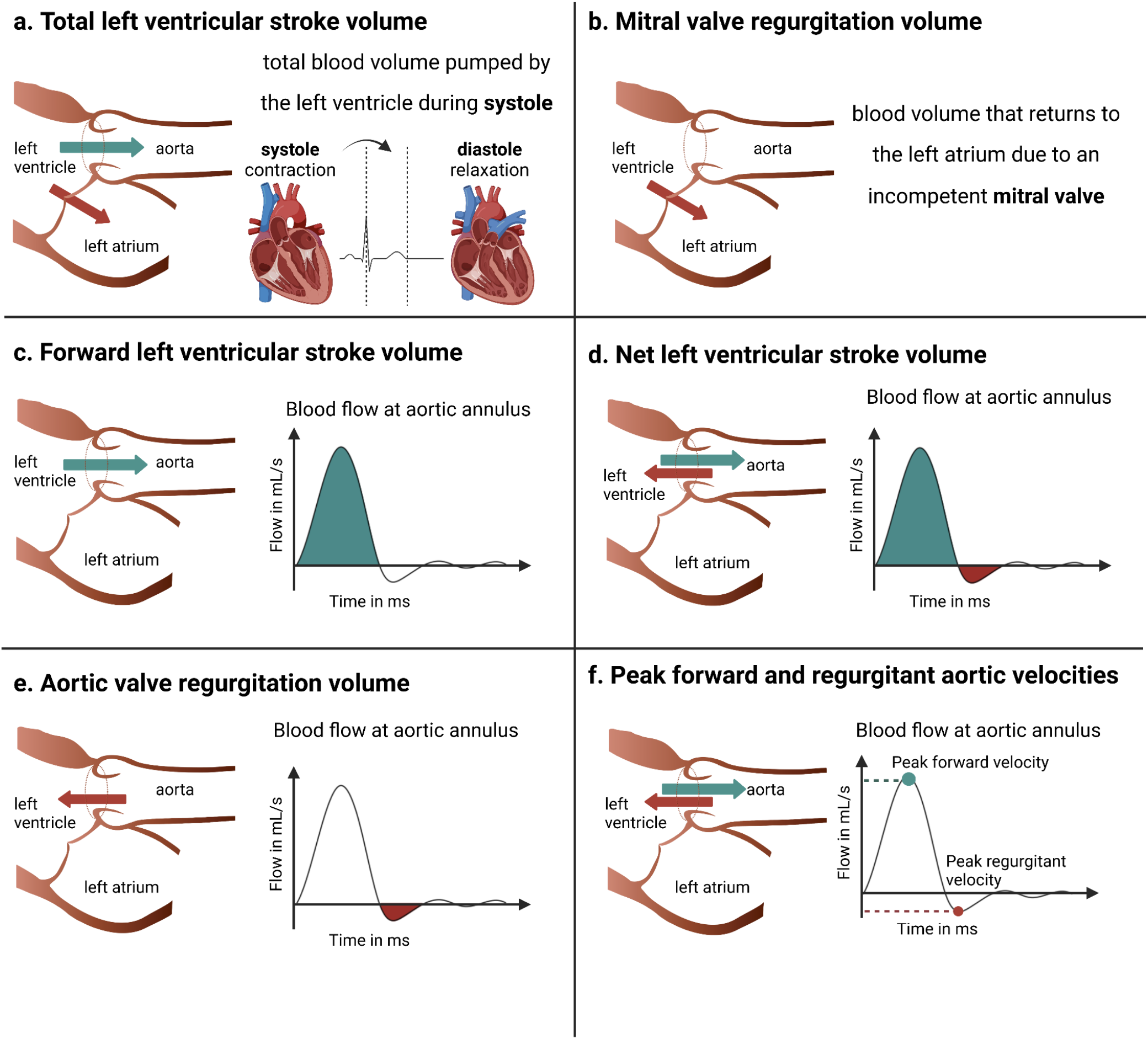
Overview of the concepts defining cardiac dynamic flow volumes analyzed in this study. Systole: left ventricular contraction phase. Diastole: left ventricular relaxation phase. **a**, Total left ventricular stroke volume is defined as the complete blood volume that is pumped towards the aorta and backwards to the left atrium during systole. **b**, Mitral valve regurgitation is the blood volume that flows backwards into the left atrium during systole due to the incompetence of the mitral valve. Also, mitral valve regurgitation can be determined by the difference between total left ventricular stroke volume and the forward left ventricular stroke volume, which is a definition commonly used for in MRI analysis^11^. **c**, Forward left ventricular stroke volume is the blood volume pumped from the left ventricle to the aorta during systole. This is equivalent to the area under the curve of the positive portion of the plot displaying aortic blood flow vs. time. **d**, Net left ventricular stroke volume is the blood volume that is effectively pumped into the aorta during an entire heart cycle: the remaining forward left ventricle stroke volume after excluding the aortic valve regurgitant volume. **e**, Aortic valve regurgitant volume is the blood volume that returns to the left ventricle during diastole due to an incompetence of the aortic valve. This is equivalent to the area under the curve of the negative portion of function plotting the aortic blood flow vs. time. **f**, The peak forward and regurgitant velocities at the aortic annulus (at the level of the sinotubular junction) correspond to the positive and negative peaks of mean aortic blood flow/velocity over a heart cycle, respectively. Of note, because over the cross-section of the aorta the velocity differs from pixel to pixel, the velocities are averaged for that aortic cross-section per time instance/frame.

In order to obtain total left ventricular stroke volume, we deployed a published algorithm^4^ on the CINE MRI dataset of the UK Biobank. For the forward and net left ventricular stroke volumes, Ao_F_Vmax and Ao_R_Vmax, aortic valve regurgitation volume and fraction, we developed DeepFlow (**Figure 2**), a deep learning-based segmentation algorithm (**Figure 3**) to achieve automated extraction of MRI phenotypes from cardiac phase contrast MRI DICOM files. We used a subset of 4,500 MRI images from 150 individuals in the UK Biobank to train this model. After training, DeepFlow’s segmentation model was tested in 450 MRI images from 15 individuals. The highest Dice similarity coefficient in the test set was 0.93, indicating good segmentation performance^12^. With this segmentation performance, the complementary characterization of the aortic annulus area (at the sinotubular junction) was enabled (see **Methods**).

**Figure 2.**
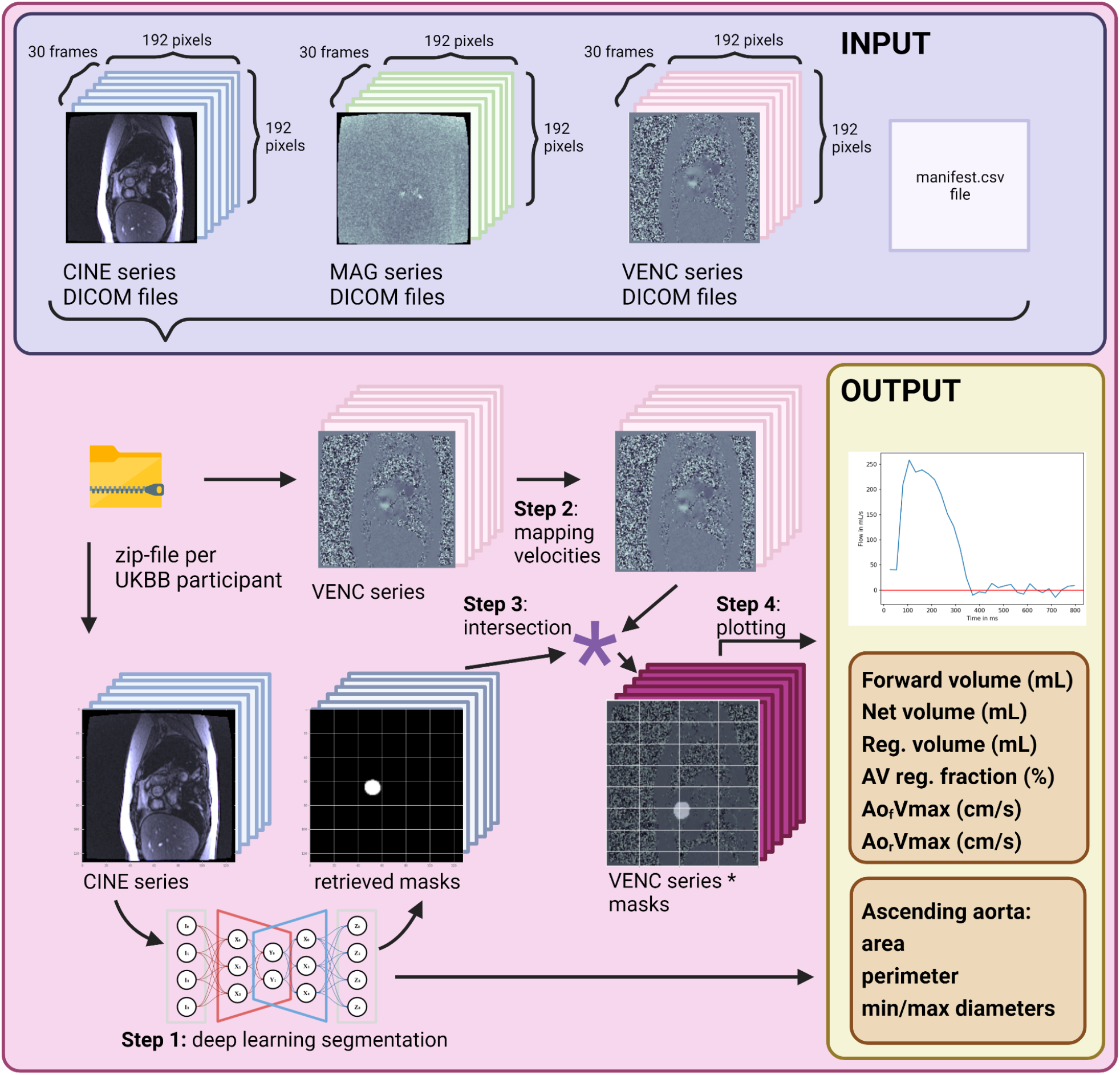
Overview of the software pipeline. The input per participant consists of 91 files (90 DICOM-files from 3 different phase contrast magnetic resonance imaging series - CINE, anatomic imaging, MAG, magnetic phase and VENC, velocity-encoded sequences - each of them with 30 frames over a heart-cycle, and one manifest file) that are compressed in a zip-file. Step 1: the software takes the DICOM-files from the CINE series and applies a deep-learning based segmentation model with pre-trained weights which retrieves the region of ascending aorta transversal at sinotubular junction (inner edge to inner edge). Step 2: the software maps the pixel-intensities of the VENC series to the corresponding velocities by applying equation 1 (for equations, see Methods). Step 3: the obtained binary masks from the CINE series are intersected with the velocity-mapped VENC series. Step 4: mean of velocities per frame are calculated and plotted over the heart cycle. Also, after applying equation 2, the sum of the volumes per frame is plotted, whereas forward volume, regurgitant volume and aortic valve regurgitation fraction (using equation 2 and 3) can be inferred. Lastly, from the masks obtained after segmentation of the CINE series, the cross-sectional area, perimeter (equation four) and minimal/maximal diameters of the ascending aorta at the sinotubular junction are returned. * denotes the multiplication/intersection operation.

**Figure 3.**
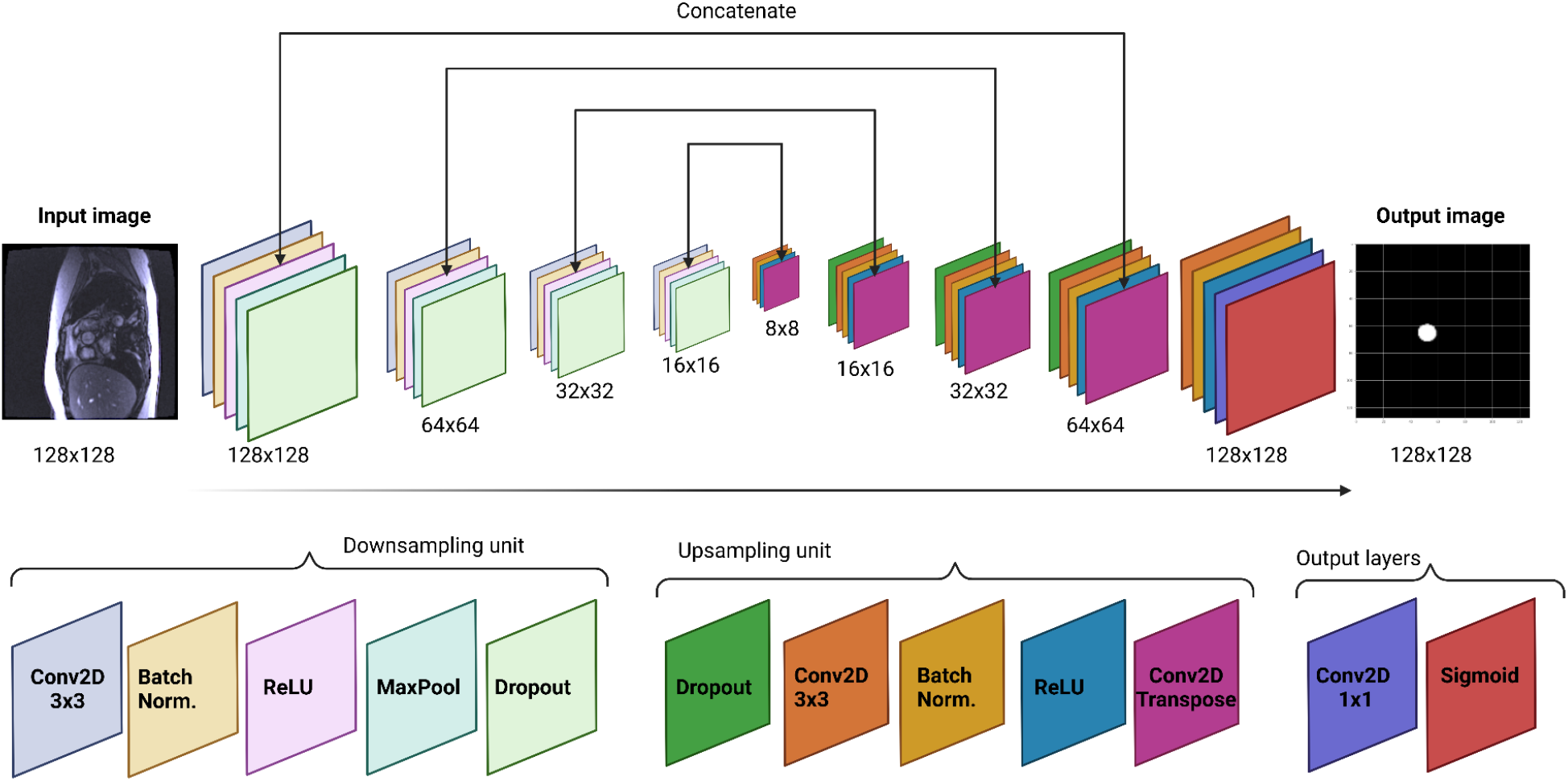
U-Net architecture of the segmentation model used in DeepFlow. The input image (128×128) is fed into a U-Net architecture, which predicts a pixelwise probability of being part of the region of interest or not. The U-Net is composed by a downsampling path and an upsampling path, which explains the u-shaped architecture. The contracting path is composed by a convolutional network that consists of repeated application of convolutions (four times), each followed by a batch normalization layer, a rectified linear unit (ReLU), a max pooling and a 5% dropout operation. The expansive pathway combines the feature and spatial information through a sequence of up-convolutions and concatenations with high-resolution features from the contracting path. Specifically, each series of up-convolution is composed by a dropout layer, followed by a convolutional operation, batch normalization, ReLU activation and a transverse convolution operation. The last layer of convolutions in the architecture consists of a convolution operation, batch normalization, ReLU activation, a last convolution operation using a 1×1 kernel and an sigmoid output layer computing the probabilistic label map (with a 128×128 size).

We tested DeepFlow on a second test set (6,000 MRI images from 200 UK Biobank participants) for which output from a commercial proprietary algorithm (Medviso^13^) was available. DeepFlow traits were highly correlated with Medviso measurements (Pearson correlation coefficients 0.96 ± 0.02 for forward left ventricular stroke volume, and 0.90 ± 0.03 for aortic valve regurgitation volume (mean± SD), **supplementary figure S2**). No significant difference was found in Medviso segment vs. DeepFlow measurements of aortic valve regurgitation volume or forward left ventricular stroke volume (aortic valve regurgitation fraction: mean difference = +0.26 mL [−3.04 - 3.56mL], p = 0.54; forward left ventricular stroke volume = −2.94 [−16.94 - 11.06] mL, p=0.11, **supplementary figure S2**).

Additionally, in order to analyze the genetic correlation with the cardiac dynamic flow volumes, we retrieved static left- and right ventricular volumes (left and right ventricular end systolic and end diastolic volumes, as well as left ventricular ejection fraction) and the measurement of left ventricular mass at an end-diastolic phase from cine imaging. All volumes, areas and left ventricular mass were indexed to Body Surface Area (BSA).

### DeepFlow enables precise measurement of aortic valve regurgitation

As additional validation, we tested the hypothesis that DeepFlow could reliably predict clinical outcomes in aortic regurgitation. In agreement with previous echocardiography studies^14–16^, our results showed that male sex and the presence of ascending aortic aneurysm were the most significant predictors of aortic valve regurgitation progression (**Figure 4**, complete results after linear regression in supplementary discussion **SD1**). Also, we found that aortic valve stenosis, hypertension, previous coronary artery bypass operations and rheumatic valve disease were significantly enriched within higher aortic valve regurgitation fraction severity groups, congruent with previous observations ^14,17^ (**Tables 1–3**). Moreover, rates of all-cause mortality (mild vs. severe: p < 0.005 and mild vs. moderate: p = 0.03, log-rank test) and new aortic valve replacement (mild vs. severe: p < 0.005 and moderate vs. severe: p = 0.02, log-rank test) were enriched in patients with higher aortic regurgitation fractions (**Figure 4**) (mild: < 22%; moderate: >= 22%^18^; severe: >= 33%^19^). Time to event analyses are included in supplementary figure **S4**.

**Table 1.** Baseline comorbidities across the aortic valve regurgitation fraction severity.

| **N = 33590** | **AR fraction** |  |  |  |
| --- | --- | --- | --- | --- |
| **Diagnosis (ICD10-code)** | **Mild**<br>**($< 22\%$)** | **Moderate**<br>**($\geq 22\% \ \& \ < 33\%$)** | **Severe**<br>**($\geq 33\%$)** | **p-value** |
| **Hypertension (I.10)** | 5501 | 174 | 89 | $3e^{-10}$ *** |
| **Diabetes mellitus**<br>**(E.10, E.11, E.12, E.13)** | 1134 | 38 | 10 | 0.76 |
| **COPD**<br>**(J.44)** | 299 | 11 | 3 | 0.95 |
| **PAD**<br>**(I.73)** | 172 | 3 | 3 | 0.92 |
| **Renal insufficiency**<br>**(N18)** | 258 | 15 | 4 | 0.06 |
| **Mitral valve disease**<br>**(I.34)** | 133 | 2 | 3 | 0.73 |
| **Tricuspid valve disease**<br>**(I.36)** | 2 | 0 | 0 | 0.99 |
| **Pulmonary valve disease**<br>**(I.37)** | 8 | 1 | 1 | 0.04 |
| **Aortic valve stenosis**<br>**(I.35.0)** | 41 | 4 | 4 | $5e^{-7}$ *** |
| **Post endocarditis**<br>**(I.33)** | 4 | 0 | 0 | 0.99 |
| **Post-myocarditis**<br>**(I.40)** | 2 | 0 | 0 | 0.99 |
| **Chronic CAD**<br>**(I.25)** | 1419 | 67 | 28 | 1e<sup>-8</sup> *** |
| **Post-MI**<br>**(I.21, I.22., I.23)** | 560 | 19 | 11 | 0.13 |
| **Atrial fibrillation or**<br>**flutter**<br>**(I.48)** | 680 | 27 | 13 | 0.17 |
| **Cardiomyopathy**<br>**(I.42, I.43)** | 59 | 4 | 2 | 0.02 |
| **Congenital heart disease**<br>**(Q.24)** | 4 | 1 | 0 | 0.36 |

ICD10-codes definitions are considered as surrogate markers of the comorbidities. \*\* indicates p-value < 0.01, \*\*\* indicates p-value < 0.0001. CAD: coronary artery disease; COPD: chronic obstructive pulmonary disease; MI: myocardial infarction; PAD: peripheral artery disease.

**Figure 4.**
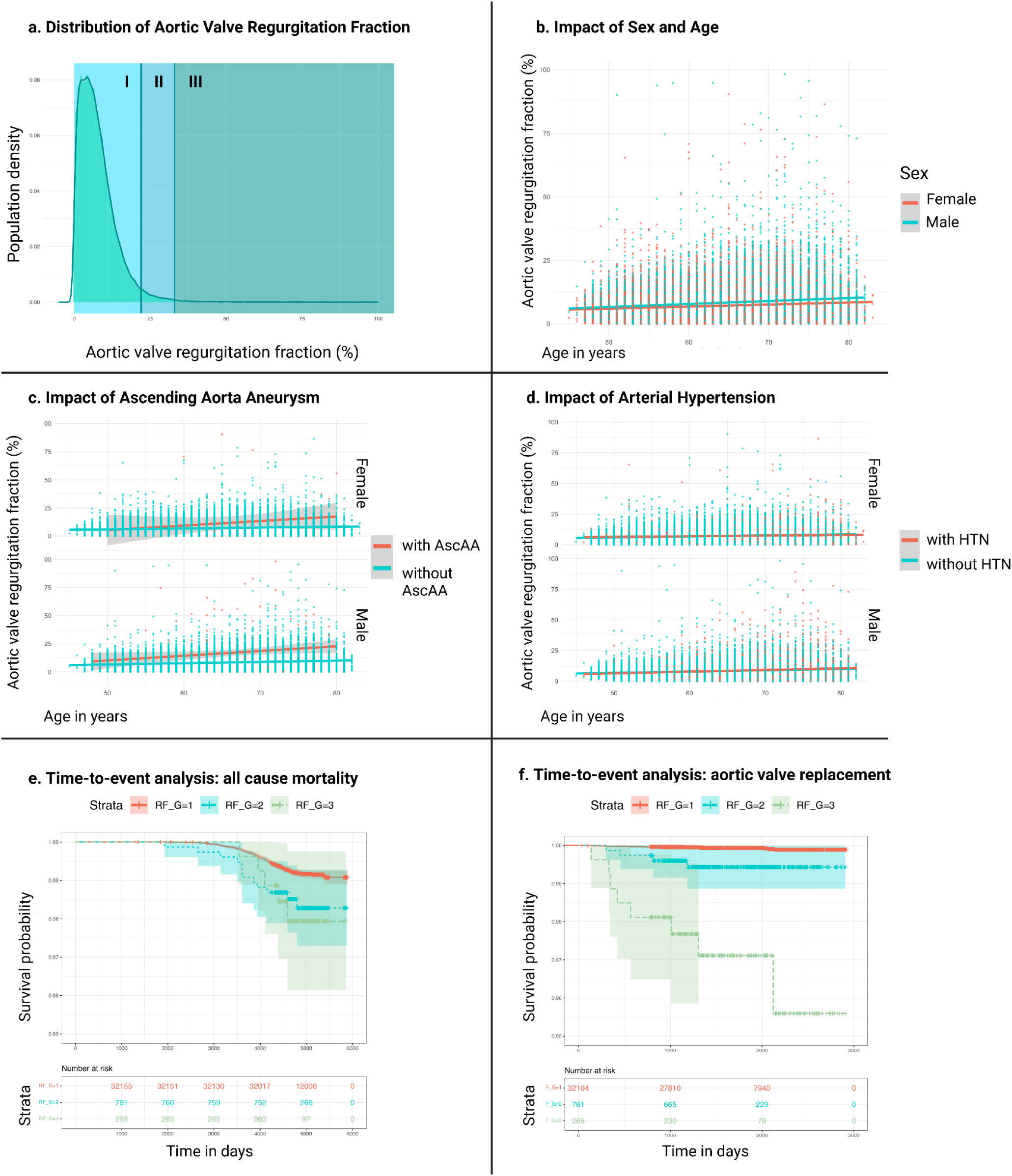
Epidemiology of aortic valve regurgitation fraction in the UK Biobank population. **a**, Density distribution histogram of the aortic valve regurgitation fraction. “I” indicates mild aortic valve regurgitation fraction (< 22%), “I” indicates moderate aortic valve regurgitation fraction (>= 22% and < 33%) and “III” indicates severe aortic valve regurgitation fraction (>= 33%). Across 40,077 UK Biobank individuals included, the mean age at the time of phase contrast magnetic resonance imaging was 64.5 ± 7.7 years, mean aortic valve regurgitation fraction was 7.7 ± 6.5 %. In order to infer the progression rates according to different risk groups we performed multivariate linear regression with aortic valve regurgitation fraction as the dependent variable. **b**, Multivariate regression analysis with sex, age at MRI visit as independent variables. Male population was found to be an independent predictor of higher AR fraction with age. **c**, Multivariate linear regression adding the presence or absence of ascending aorta aneurysm as an independent variable, which was a significant predictor of higher aortic valve regurgitation fraction with age. **d**, Multivariate linear regression considering the presence or absence of hypertension as an independent variable, which was not a significant predictor of higher aortic valve regurgitation fraction. Linear regression fitting equations presented in **supplementary equations**. For the time-to-event analysis of **e**, all-cause mortality and **f**, new aortic valve replacement, common concomitant cardiovascular comorbidities were excluded (see Methods). Also, aortic valve regurgitation fraction was adjusted to sex and age at MRI. Log-rank test was performed to assess statistical significant differences between the incidence of outcomes. The percentiles of 99.2% for severe and 96.9% for moderate aortic valve regurgitation fraction were used to define the groups. RF_G indicates aortic valve regurgitation fraction severity group: red: mild (1); blue: moderate (2); green: severe (3). AscAA indicates aneurysm of the ascending aorta; HTN indicates hypertension.

### Aortic valve regurgitation has genetic underpinnings distinct from left ventricular volumes

We next investigated the underlying genetic architecture of cardiac dynamic flow volumes. For this purpose, we used DeepFlow-derived traits, as well as left and right ventricular parameters^4^ and mitral valve regurgitation volume. We first examined heritability (h^2^, **Figure 5 A**) using the linkage-disequilibrium (LD) regression score^20^ and found that structural phenotypes, aortic annulus area, and BSA-indexed left ventricular mass were the two most heritable traits (h^2^= 0.40 ± 0.04 and 0.36 ± 0.04, respectively). Aortic valve regurgitation fraction and aortic valve regurgitant volume showed low heritability (h^2^ =0.06 ± 0.02 and 0.07 ± 0.02, respectively). Mitral valve regurgitation volume was associated with the lowest heritability (h^2^ =0.03).

**Figure 5.**
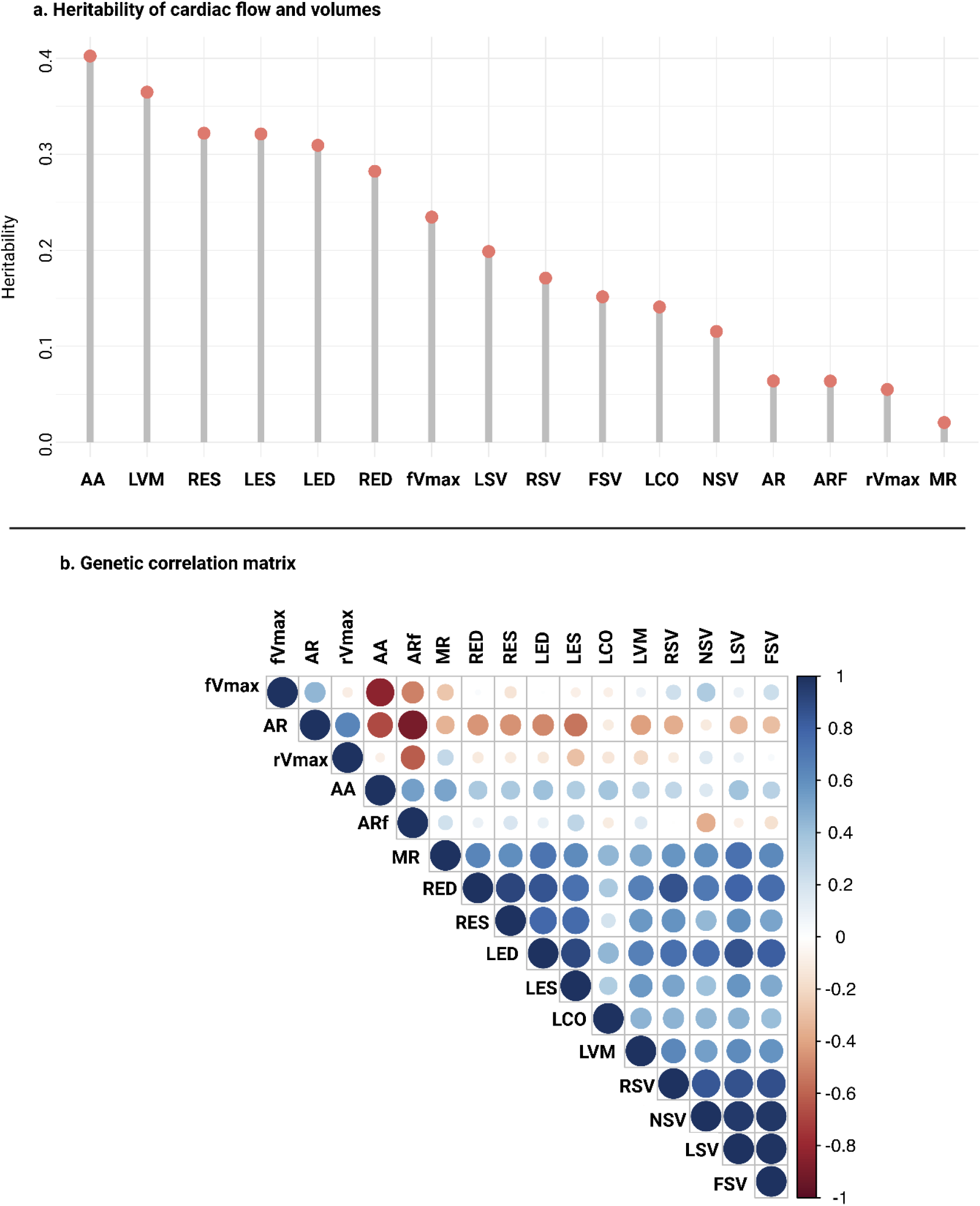
Heritability and genetic correlation matrix of cardiac flow dependent variables. **a**, Heritability of the cardiac flow and volume parameters, ranked from trait with highest heritability to lowest. **b**, In the genetic correlation matrix, the size of the dots corresponds to the magnitude of the correlation between variables. The color indicates the direction of the correlation (blue end: positive correlation, red end: negative correlation, white: no correlation). AA indicates: BSA-indexed aortic annulus area; AR: BSA-indexed aortic valve regurgitant volume; ARf: aortic valve regurgitation fraction; fVmax: forward peak velocity at aortic annulus; FSV: BSA-indexed forward left ventricular stroke volume; LCO: BSA-indexed left cardiac output; LSV: BSA-indexed left ventricular stroke volume; MR: BSA-indexed mitral valve regurgitation volume; NSV: BSA-indexed net left ventricular stroke volume; RES: BSA-indexed right ventricular end systolic volume; REV: BSA-indexed right ventricular end diastolic volume; RSV: BSA-indexed right ventricular stroke volume; rVmax: aortic retrograde peak velocity.

We studied the genetic correlation between these traits using the LD regression score (rg, **Figure 5 B**). Aortic valve regurgitation parameters (aortic valve regurgitation fraction and volume) were highly correlated with each other (rg = −0.91 ± 0.06, p-value = 5.7e^−55^), as well with peak regurgitant velocity (Ao_R_Vmax) (rg = −0.61 ± 0.1, p-value = 1.7e^−6^). Moreover, aortic valve regurgitation fraction was positively genetically correlated with aortic annulus area (Ao_F_Vmax) (rg = 0.53 ± 0.1, p-value = 8.4e^−7^) and negatively correlated with aortic peak forward velocity (Ao_F_Vmax) (rg −0.51 ± 0.1, p-value = 2.2e^−6^). There was a negative genetic correlation between the aortic annulus area and Ao_F_Vmax (rg −0.83 ± 0.04, p-value = 2.5e^−111^). This is in alignment with Bernoulli’s principle, according to which fluid velocity is inversely associated with cross sectional area. Aortic valve regurgitation fraction displayed weak genetic correlations with both left and right ventricular volumes (rg < |±0.4| ± 0.13). This was in contrast with mitral valve regurgitation volume, which was more strongly and positively genetically correlated with left and right ventricular volumes (rg > |±0.5| ± 0.37).

### Novel genome-wide loci are associated with dynamic flow volumes

We next sought to identify common genetic variants that were associated with DeepFlow derived phenotypes (**Figure 6**). We computed LD-regression score intercepts and genomic control lambdas to assess for genomic inflation in our GWAS (supplementary figure **S5**), which were acceptable. For BSA-indexed aortic annulus area, 56 distinct loci were identified (Supplementary table **S6**), validating 27 loci previously reported in association with CINE MRI-derived aortic areas^12^ (**Figure 6 B)**. Leading SNVs for this trait were found to be in close proximity to *ELN* (−log_10_ p= 52.75), as well as *PRDM6* (−log_10_ p = 22.94) and *FBN1* (−log_10_ p = 22.25), all of which are genes associated with Mendelian aortic diseases. Forty-one genome-wide significant loci were associated with peak forward velocity at the aortic annulus (Ao_F_Vmax) (**Figure 6 A**, Supplementary table **S7)**, with the most significant single nucleotide variant (SNV, rs11768878) also near the elastin gene *ELN* (−log_10_ p = 43.30). The lead SNV in locus *LINC01808* (long intergenic non-protein coding RNA) for aortic valve regurgitation fraction (−log_10_ p= 7.89, Supplementary table **S10**) was also significantly associated with Ao_F_Vmax and aortic annulus area (−log_10_ p = 18.07 and 15.97, respectively), demonstrating internal validation of our findings. Overall, Ao_F_Vmax and aortic annulus area shared 35 significant loci.

**Figure 6.**
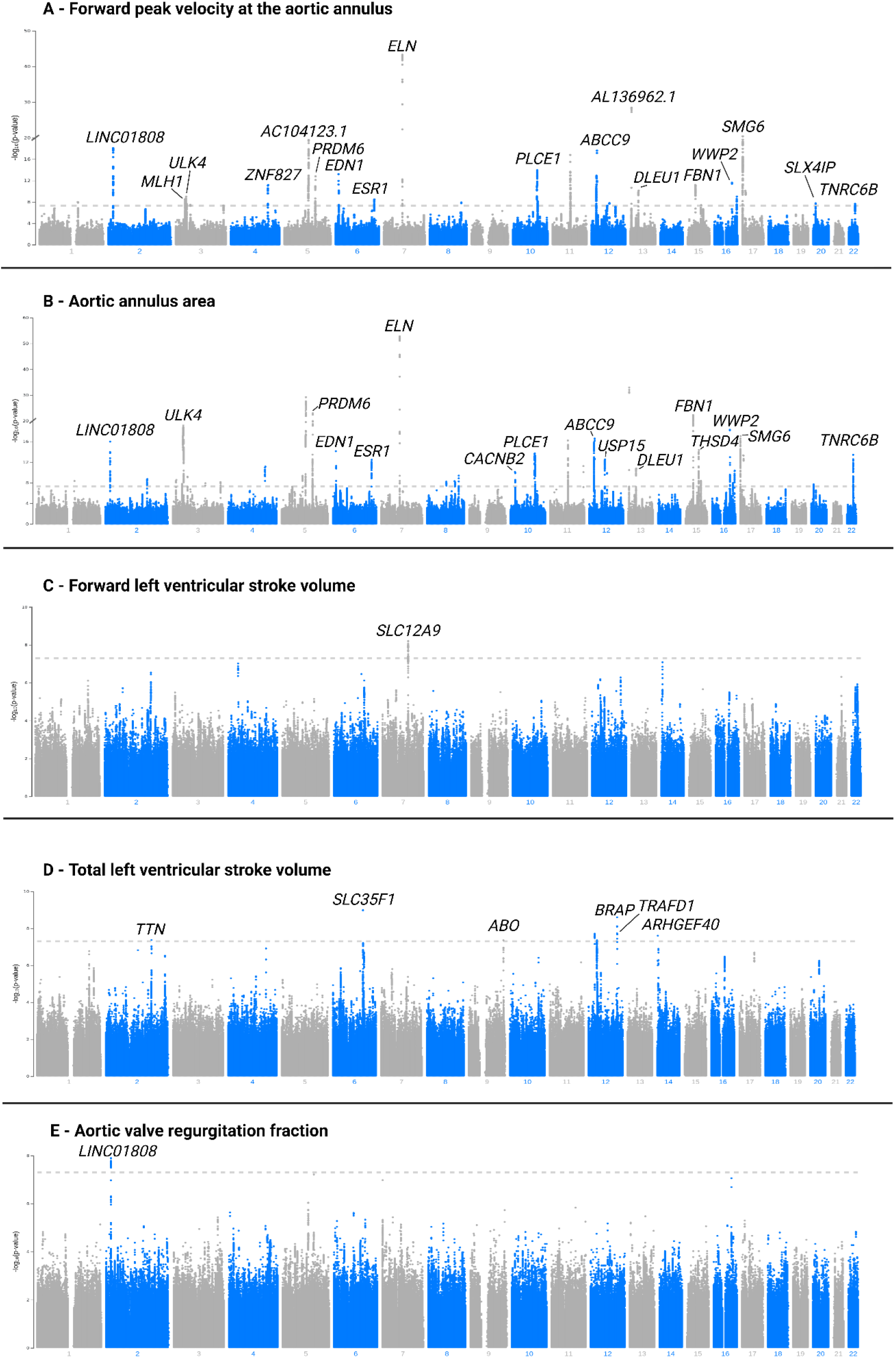
Manhattan plots showing the GWAS results of **a**, aortic valve regurgitation fraction; **b**, peak forward velocity at the aortic annulus; **c**, aortic annulus area; **d**, forward left ventricular stroke volume; **e**, total left ventricular stroke volume. All patients with phase contrast magnetic resonance imaging and imputed microarray DNA data. Independent variables were adjusted to age, sex, the first 5 genetic principal components and genotyping array. To adjust for multiple testing, we applied Bonferroni correction using the genome-wide significance threshold of P < 5 × 10^−8^ for selection of independent SNVs that were considered significantly associated with the traits. Complete lists of the genomic loci are shown in supplementary tables S6-S11. All volumes and areas are BSA-indexed.

With respect to dynamic left ventricular volumes, we found one SNV (rs34098933) near *SLC12A9* that was significantly correlated with forward left ventricular stroke volume (−log_10_ p = 8.33, Supplementary table **S11**). Eight distinct loci were identified for total left ventricular stroke volume, whereas SNVs closest to *SLC35F1* (−log_10_ p = 8.99) and *BRAP* (−log_10_ p =8.60) loci showed the most significant association (Supplementary table **S8**). The *SLC35F1* (solute carrier family 35 member F1) gene, enabling transmembrane transporter activity, has been linked to the PR-interval of the electrocardiogram^21^, but has not yet been linked to this trait in previous studies ^9,22^. On the other hand, the *BRAP* gene, which has been recently implicated as a regulator of the cardiomyocyte cell cycle^23,24^, was previously reported in association with total left ventricular stroke volume in two prior studies^9,22^. Our GWAS for Ao_R_Vmax, BSA-indexed net left ventricular stroke volume and BSA-indexed mitral valve regurgitation volume did not reveal any significant associations (respective Manhattan plots included in Supplementary figure **S7)**. GWAS for BSA-indexed aortic valve regurgitation volume (Supplementary figure **S7**) revealed two lead SNVs associated with this trait (rs77097530 (−log_10_ p = 7.48) and rs4719607 (−log_10_ p = 7.34), Supplementary table **S9**). SNV rs77097530 is closest to protein coding downstream gene *CEP120* and upstream gene *PRDM6*, while the latter has been linked to both bicuspid aortic valve and aortic root disease.

Elimination of participants with common cardiovascular comorbidities (as performed in the time-to-event analysis) from the population set did not reveal additional significant SNVs, and narrowed the number of loci significantly associated with Ao_F_Vmax and aortic annulus area. However, this secondary analysis showed an additional novel locus (*GNA12*/*AMZ1*, encoding a GTPase and a metallopeptidase, respectively) associated with both aortic valve regurgitation fraction (−log_10_ p = 7.70) and volume (−log_10_ p = 7.66) (Supplementary discussion **SD2**, flowchart of the GWAS sample size in supplementary discussion **SD3**).

### Genetic determinants of aortic root size and flow metrics map to connective tissue and blood pressure pathways

We next identified potential interactors of the genes closest to loci significantly associated with Ao_F_Vmax and aortic annulus area. We performed unsupervised gene clustering using K-means followed by enrichment analysis for pathways and phenotypes within identified clusters using the STRING-DB server^25^. We took into account the most significantly predicted phenotypes from Gene Ontology^26^, the Monarch Human Phenotype Ontology (HPO)^27^ and UniProt^28^. For aortic annulus area, there were three major gene clusters identified. Genes in these clusters were highly associated with pulse pressure management (Monarch HPO 0005763, q-value = 2.6e^−6^), diastolic blood pressure (Monarch HPO 0004324, q-value = 9.1e^−5^) and William-Beurer syndrome (a condition caused by a microdeletion involving *ELN* and *LIMK1*)^29^, (q-value = 0.04), respectively. In the gene cluster most highly associated with diastolic blood pressure, extracellular matrix genes (UniProt KW-0272, q-value = 0.02) and aortic root size-associated genes (Monarch HPO 0005037, q-value = 0.03), were also highly enriched. For Ao_F_Vmax, two major clusters were defined: The first was most significantly enriched for microfibril-associated genes (GO 0001527, q-value = 0.03), which are related to connective tissue phenotypes, and the other was enriched for genes associated with systolic blood pressure (Monarch HPO 0006335, q-value = 0.01) (**Figure 7 A and B**).

**Figure 7.**
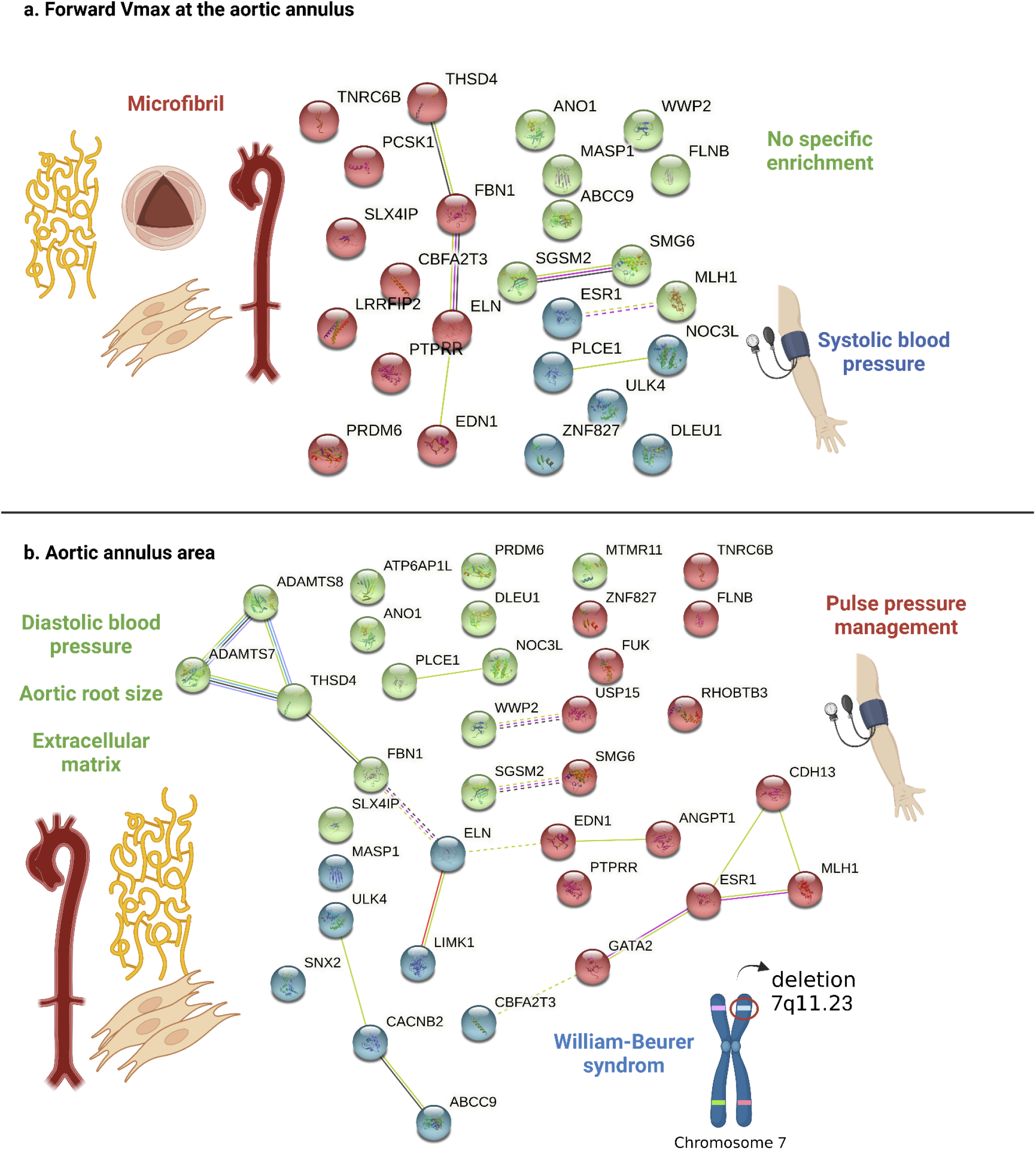
Cluster groups and gene interactions of the traits **(a)** forward peak velocity at the aortic annulus and **(b)** aortic annulus area. K-means was applied to define the three groups. In **a**, the color red represents the cluster of genes which most significantly enriched for microfibril-associated genes; color blue represents a group of phenotypes which was most significantly associated with systolic blood pressure; in color green the cluster of genes did not show a specific enrichment. In **b**, the color red represents a group of genes related which was most significantly associated with pulse pressure management; in blue, the group of genes were most significantly linked to the William-Beurer syndrome; the color green represents a cluster of phenotypes which was most significantly enriched for diastolic blood pressure, but also includes traits related to connective tissue. Complete list of phenotypes included in the clusters available in the supplementary data. Edges show protein-protein relationships, i.e., when protein work together to perform a common task. Light blue line: known interactions from curated datasets; purple line: experimentally determined known interactions; dark blue line: predicted interactions from gene neighborhood; red line: predicted interactions from gene fusions; dark blue line: predicted interactions from gene co-occurrences; light green line: predicted interaction from text mining; black line: co-expression; light purple: gene homology.

To identify tissues enriched for expression of genes near trait-associated loci, we used DEPICT^30,31^. No enriched gene sets were identified. However, tissue enrichment analysis revealed that expression of genes nearest loci associated with BSA-indexed aortic annulus area were enriched in arterial tissue (led by MeSH term A07.231.114, p-value = 7e^−4^, FDR <0.01). These results are consistent with the hypothesis that genetic drivers of aortic regurgitation act within connective tissue to affect structural traits such as aortic annulus size. Genes near loci associated with Ao_F_Vmax were significantly enriched in the myometrium (the smooth muscle of the uterus, led by MeSH term A05.360.319.679.690, p-value = 0.002, FDR <0.05). This is a surprising finding that has been previously reported in association with aortic areas derived from CINE sequences^12^. It may reflect a sex-specific association of aortic dilation, and requires additional investigation to understand its implications. Aortic valve regurgitation fraction associated loci were not significantly enriched in any tissue (Supplementary figure **S8**).

### Aortic root size is causally related to aortic valve regurgitation fraction

We next used Mendelian randomization (MR) to assess the causal relationships between aortic annulus area and aortic valve regurgitation fraction. Using the CAUSE algorithm, which better addresses horizontal pleiotropic effects compared to other methods, we observed that BSA-indexed aortic annulus area was causally associated with aortic valve regurgitation (p=0.004, **Figure 8A**). This causal interaction was also identified by applying CAUSE and three additional (two-sample) MR methods on non-overlapping random subsets of the patients (inverse variance weighting, MR-Egger, and weighted median estimation, see Methods and **Table 4**). When treating aortic valve regurgitation as the exposure (i.e., testing for the reverse causal relationship), the CAUSE analysis was not significant (p=0.11). The unidirectionality was supported by the MR-Steiger directionality test (correct causal direction is true, p-value = 3.0e^−119^).

**Figure 8.**
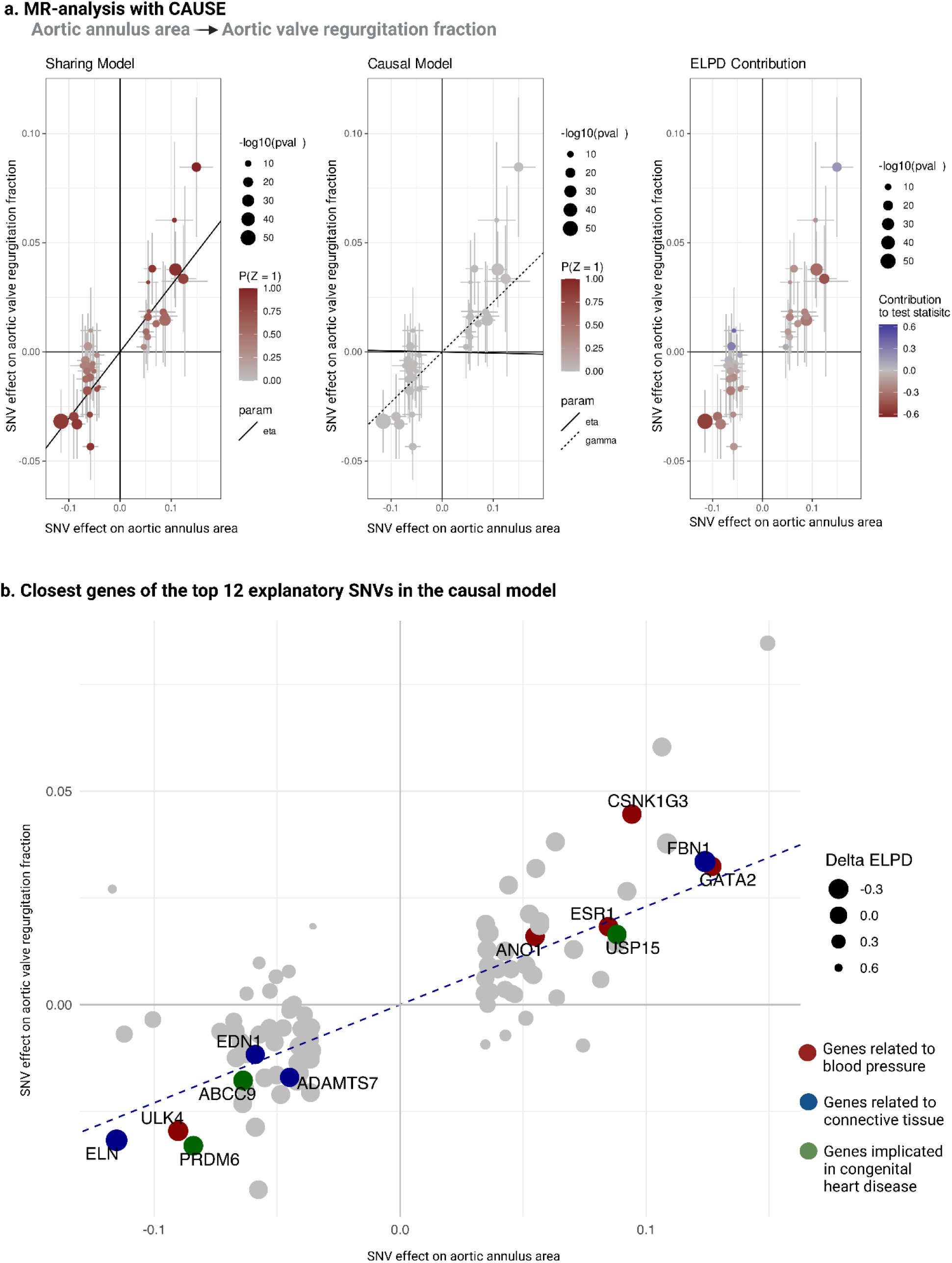
Mendelian randomization results using CAUSE to establish causal relationships between aortic annulus area (exposure) and aortic valve regurgitation fraction (outcome). As instrumental factors for each of these traits, we considered all SNVs with a P-value threshold of 5 * 10^−5^ as exposure. **a**, Scatter plots showing the SNV effects on the exposure and outcome traits according to the sharing model (scatter plot on the left) and the causal model (middle scatter plot). The continuous and dashed lines correspond to the linear fit in the sharing and causal models, respectively. The eta and gamma parameters represent the slope of the linear fit. After applying CAUSE, which uses a Bayesian model comparison approach, the causal model showed the best fit. The size of the dots in the scatter plot are proportional to the value of the SNV’s −log_10_ p-value associated with the exposure factor. We used the estimated difference in the expected log pointwise posterior density (delta ELPD), as a measurement of the explanatory value of the SNVs included in the CAUSE models. In the left scatter plot, the delta ELPD values of the SNVs are color mapped from the extreme negative (red) to the extreme positive (blue). SNVs with reddish shades favor the causal model. **b**, Subset of the causal model scatter plot, highlighting the closest genes to the top 12 SNVs with the highest negative delta ELPD values. The size of the dot associated with each SNV is proportional to the value of delta ELPD. The genes highlighted have been found to be associated with connective tissue, blood pressure management and congenital heart disease traits (coded in blue, red and green, respectively). Delta ELPD: estimated difference in the expected log pointwise posterior density; SNV indicates single nucleotide variant; Z-Score associated with the exposure related SNV’s p-value.

CAUSE uses a Bayesian model comparison approach that estimates how well the posterior distributions of a particular model are expected to predict a new set data. In the process, it computes variant-level importance scores, the difference in the expected log pointwise posterior density^32^ (delta ELPD), in which the most negative score favors the causal model. We therefore used these scores to rank the variants by their explanatory value (**Figure 8B**). For the causal model with aortic annulus area as the exposure and aortic valve regurgitation fraction set as the outcome, the three most significant SNVs included rs7782306 (closest to *ELN*), rs1036476 (intronic variant in *FBN1*) and rs61744388 (missense variant *ULK4*). The *ELN* and *FBN1* genes are implicated in connective tissue related pathways^12^. Therefore, these genes are expected to mechanistically affect the aortic annulus area, and are not expected to be horizontally pleiotropic with aortic valve regurgitation. Indeed, as can be seen in Figure 8B, the causal estimates of their variants are proximal to the MR slope, corroborating our causal estimation results (as these variants are more plausible to be proper instrumental variables based on the substantive prior knowledge). As an additional candidate to be a proper instrument, we identify *ULK4*, which has been linked to blood pressure management pathways^33^.

## Discussion

Here, we present three advances. First, we introduce DeepFlow, an open source solution for accurately capturing cardiac dynamic flow volumes from phase-contrast cardiac MRI data at scale. Second, genome-wide association studies of cardiac flow dynamic traits reveal multiple significant new and replicated loci. Third, we report a causal relationship between aortic root size and aortic valve regurgitation fraction.

Our findings have several implications. First, GWAS revealed several novel genomic loci associated with cardiac flow dynamic volumes. Interestingly, the only variant significantly associated with forward left ventricular stroke volume mapped to SLC12A9, a gene coding a solute carrier responsible for potassium/chloride symporter activity and found to be associated with the electrocardiogram RR interval. This implies a mechanism consistent with the Frank-Starling relationship^34,35^ whereby cardiac muscle responds to stretch with lengthening (increasing ventricular volume) and a more forceful contraction. A similar mechanism may also explain the association between total left ventricular stroke volume and the variant mapping to ARHGEF40. This gene is also associated with the PR interval of the electrocardiogram ^36,37^ suggesting that longer diastolic filling leads to a larger stroke volume through the Frank-Starling mechanism. That there was no overlap between the loci significantly associated with total left ventricular stroke volume and those significantly associated with aortic valve regurgitation fraction and peak forward velocity at the aortic annulus reinforces the argument that aortic valve structure and not left ventricular function or size primarily determine aortic valve regurgitation.

Mendelian randomization demonstrates a unidirectional causal relationship between aortic root size and aortic valve regurgitation. While these two findings often co-exist in patients, the causal nature of their relationship has been less clear until now. The two most explanatory variants map to *ELN* and *FBN1*, genes linked to the mendelian conditions Ehlers-Danlos syndrome^38^ and Marfan syndrome,^39^ which cause aortic enlargement. The third most explanatory variant was rs61744388, which overlaps the gene *ULK4*. Although *ULK4* encodes a member of the unc-51-like serine/threonine kinase family with a primary role in neuronal growth and endocytosis^40^, not only have prior studies suggested an association with blood pressure phenotypes^41^, they have also linked this gene to acute aortic dissection^42^. Similarly, *ABCC9, ANO1* and *PRDM6* (genes mapped by variants in the top 12 ranked explanatory variables), have been also shown to be significantly associated with the risk of acute aortic dissection^43^. *ANO1* plays a major role in regulating chloride and calcium currents in smooth muscle cells and knockout of this gene lowers vascular tone^44^. Overlap between genetic associations of the anatomy of the aorta and the physiology of blood flow, as well as the causal relationships we have uncovered, suggest that therapies used to prevent progression of ascending aortic aneurysm^45^ may help prevent progression of aortic regurgitation itself. They further suggest that components of the extracellular matrix specifically control this phenomenon, and that targeting blood pressure with agents that modify its fibrous composition (e.g., ACE-inhibitors) may be an effective strategy.

Our study has limitations. Since we did not manually curate every imaging study, instead relying on a semi-automated accurate deep learning model, some error may be introduced. To minimize noise, we set a threshold of phenotype values more than three interquartile ranges above the first quartile or below the third quartile (Winsorization method). In addition, our model was not trained to detect bicuspid aortic valves. However, this is unlikely to be a major factor given its low estimated prevalence in this population^46^. Lastly, the UK biobank is a population of predominantly European ancestry, limiting the extension of our findings to other populations.

This report demonstrates the breadth of inquiry made possible by automated phenotype extraction from population imaging databases. While dynamic flow volumes are central to health and cardiac disease diagnosis including valvular insufficiency, automated extraction of blood flow patterns to other organs (e.g., the brain) may enable discovery of genetic underpinnings of poorly understood diseases across diverse physiological systems. Further, the specific hypotheses we raise here regarding the genetic contributions of connective tissue to aortic annulus size and aortic regurgitation motivates further investigation into the prevention of aortic regurgitation progression in prevalent disease, potentially with preexisting therapeutics.

## Data Availability

All data produced in the present study are available upon reasonable request to the authors.

## URLs

**Ukbb_cardiac** https://github.com/baiwenjia/ukbb_cardiac, **CAUSE** https://jean997.github.io/cause/, **DEPICT** https://github.com/perslab/depict, **LocusZoom** http://locuszoom.org/, **SAIGE** https://github.com/weizhouUMICH/SAIGE, **STRING-DB** https://string-db.org/, **TwoSampleMR** https://github.com/MRCIEU/TwoSampleMR.

## Acknowledgments

This research has been conducted using the UK Biobank Resource under Application Numbers 63735 and 22282. Figures were created with BioRender.com and LocusZoom.org. BG received funding from the Deutsche Forschungsgemeinschaft (DFG – German Research Foundation) under the Walter-Benjamin Program (GO 3196/3-1, 707766 - 809341).

## Author Information

### Contributions

B.G. and E.A. designed the study. B.G. trained, validated and tested DeepFlow with contribution of A.S.; conducted the GWAS and epidemiology related analyses; wrote the supplementary material and tables. B.G designed the figures with the contribution of A.S.. B.G., E.A. and V.P. wrote the manuscript with contributions of A.S., D.A., J.S., M.S. and F.H.. B.M. gave advice in the conception of the project. A.S. performed software implementation of DeepFlow, as well as ensured cross-platform compatibility. A.S. contributed to the time-to-event analyses of clinical outcomes in aortic valve regurgitation. D.A. contributed and advised the Mendelian Randomization analysis.

## Methods

### Derivation of flow dependent variables from phase contrast MRI

The UK Biobank is a prospective study of > 500,000 individuals from the UK enrolled between 2006-2010 with extensive phenotype, imaging and multiple genetic characterizations, which has been previously described ^47,48^. The UK Biobank received ethical approval from the National Health Service’s National Research Ethics Service North West (11/NW/0382).

The latest release of UK Biobank’s MRI dataset comprises data from > 40,000 participants. The MRI dataset contains eight cardiac imaging sets, including CINE sequences of long- and short-axis views of the left ventricle, the Shortened MOdified Look-Locker Inversion recovery (shMOLLI) sequences, aortic distensibility and left ventricular outflow tract imaging, CINE tagging and scout, and phase contrast magnetic resonance imaging sequences. All MRIs were taken using the Siemens scanner equipment (Siemens, Munich, Germany) and the images are stored in DICOM format. The phase contrast magnetic resonance imaging sequence is crucial to obtain aortic blood flow related variables.

There are three series of phase contrast magnetic resonance imaging of the aortic valve registered in an *en face* view at the sinotubular junction: raw anatomical images (CINE), magnitude (MAG) and velocity encoded (VENC) (**Figure 2**). While pixel-value MAG and VENC series can be mapped to the real velocity of blood-flow at that specific pixel-coordinate, the pixel-values of the CINE series are unrelated to velocity information. For the Siemens scanner, equation one maps the pixel-intensity value from the VENC series to the corresponding velocity in cm/s.

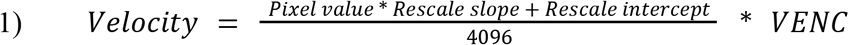

In equation one, rescale intercept and rescale slope information can be obtained from the (0028,1052) and (0028,1053) DICOM-tags metadata, respectively. The VENC (largely set to 250 cm/s in the UK Biobank) parameter can be obtained from the CSA headers, which are Siemens-specific private data elements embedded in DICOM headers. According to the UK Biobank MRI study protocol^10^, each of these phase contrast magnetic resonance imaging series (CINE, MAG and VENC) are composed of 30 frames taken over a heart cycle with each frame corresponding to 12-bit grayscale images, with a matrix-size of 192 × 192 pixels. Additionally, one manifest-file per phase contrast magnetic resonance imaging is provided by the UK Biobank. Each participant has, therefore, a total of 90 DICOM- and one manifest-files, compressed into a single compressed ZIP archive. Phase contrast magnetic resonance imaging series that diverged from the protocol were excluded from the analysis.

Considering the 30 frames composing the VENC series of the phase contrast magnetic resonance imaging sequence, if the region or mask of the aortic annulus can be segmented, the images obtained after intersection between the predicted masks and the velocity corrected VENC imaging series deliver the blood flow velocity at each pixel coordinate. Here, aortic annulus is defined as the transversal section of the ascending aorta at the sinotubular junction. Regarding the collection of mean velocities per frame at the sinotubular region ([sum of velocities of aortic annulus area]/[number of pixels in aortic annulus area]), the forward peak velocity (Ao_f_Vmax) and regurgitant peak velocity (Ao_R_Vmax) correspond to the highest and lowest mean velocities of this collection, respectively. Equation two calculates the total net left ventricular stroke volume calculation over an entire heart cycle using the pixel-intensity-values after applying equation one^49^.

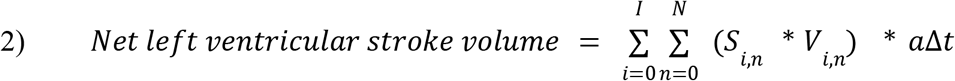

In equation two, *i* stands for the segment index, *I* for the number of temporal instances in the scan, *N* for the number of pixels in each segment, *S* for the binary segmentation map, *V* for the velocity map calculated using equation 1, *a* for the pixel area (in cm^2^), and finally, Δt for the time interval between segments. Pixel area and time interval are retrieved using the (0028,0030) and the (0018,1063) DICOM-tags, respectively. Next, volumes per frame obtained in equation two can be plotted over a heart cycle, and both regurgitant and forward volumes are calculated as the sum of all negative and positive volumes, respectively. Equation three delivers the definition of aortic valve regurgitation fraction, the main target phenotype feature.

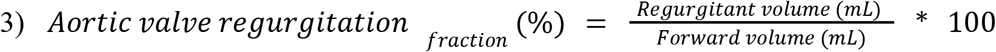

Furthermore, using the predicted binary masks, the maximal/minimal areas, diameters and perimeters can be computed. The area is calculated as the sum of all pixels-values. The maximal and minimal diameters are retrieved after calculating the centroid of the mask and subsequent bounding box’s height and length. Finally, the perimeter can be obtained by assuming the shape of the aorta’s transversal plane as an ellipse and applying the Ramanujian’s equation shown in equation four.

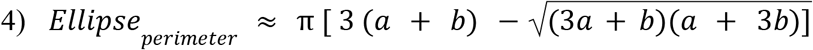

In equation four, *a* corresponds to the major axis and *b* to the minor axis. For the subsequent analyses in this study, we used the maximal area computed over a heart cycle. With these considerations, the main computational steps involved in the described software were derived and are depicted in **Figure 2**.

### Preprocessing and deep learning-based segmentation

In order to compute the model segmentation’s weights, an autoencoder deep learning algorithm, U-net, was used. U-nets are powerful for image segmentation tasks. The model architecture is depicted in **Figure 3** and can be accessed in the GitHub repository: https://github.com/Urban90/deepFlow/blob/master/assets/model.svg. Our U-Net architecture is characterized by several convolutional layers that are used to extract image features. A 3×3 kernel is used for each convolution, which is followed by batch normalization, an activation layer with a rectified linear unit (ReLU) and a dropout layer (set to 5%). Then, the feature map is downscaled by a factor of 2 after every convolution in order to learn features at a more global level. Afterwards, it iteratively up-samples the feature map stepwise to its original resolution by a factor of 2 using transposed convolutions and concatenates with the feature map at the next scale. Downsampling and upsampling part of the U-Net consists of four convolutional layers, respectively. The last element of this U-Net architecture includes a further convolutional layer with a 3×3 kernel, followed by batch normalization, ReLU activation layer, a last convolutional layer with a 1×1 kernel and a sigmoid output layer computing a probabilistic label map. Input and output images have the shape of 128 x128 pixels. Details regarding the hyperparameters of the convolutional and batch normalization layers are compiled in **Supplementary tables 1 to 3**. Training, validation and testing of the software was performed with Python version 3.9.7, with module tensorflow-gpu version 2.4.1.

In the preprocessing steps, first, manual segmentation of the complete CINE series from 150 randomly selected participants (30 frames per participant, conferring a total of 4,500 frames) was performed by a level III MRI trained physician (BG) using Medviso’s open-source software Segment^13^. Then, the manually segmented images were then converted into a binary mask and the pixel intensities corresponding to the region of interest were set to 1 (excluding the edge of the segmentation line). The remaining pixels were set to 0. Second, the pairs composed by the CINE series and the respective binary mask were resized to 128 × 128 pixels and checked for grayscale scaling (pixel-intensity ranging from 0 to 1 in the masks produced for training). After this step, training and validation subsets were randomly produced following a 9:1 ratio, and was done over 10 epochs and a batch size of 4. The model was compiled with the “Adam” optimizer. Per training epoch loss quantification was calculated using mean squared error. We also defined callback functions to automate the following tasks after every epoch: stopping training after 5 epochs with no improvement, adjusting the learning rates over time by 0.1 factor (with a minimum learning rate of 0.00001) if the loss metric did not improve after 5 epochs.

For the test set, manual segmentation of 300 CINE images from the phase contrast magnetic resonance imaging sequences of 15 randomly selected patients was also performed by a level III MRI trained physician (BG) following the same steps as previously described. After thresholding the masks’ predicted pixel intensities to either 1 if the pixel value was higher than 0.5 or to 0 otherwise, the Dice similarity coefficient was calculated. Lastly, the best model weights are saved to be deployed in the segmentation prediction of the remaining studies in the phase contrast magnetic resonance imaging dataset.

### Postprocessing computation

After segmentation, in order to compute the aortic blood flow dependent variables, the first step includes mapping of the pixel intensities of the VENC imaging series to the real velocity (cm/s) using the previously described equation one. The mathematical operation enunciated in its numerator (*pixel value* * *Rescale slope*) + *Rescale slope*) is automatically performed through the DICOM-file conversion to NIfTI-format (from the Neuroimaging Informatics Technology Initiative, which can also be applied to cardiovascular imaging)^50^. Pixel-wise multiplication by factor *VENC*/4096 of the resulting image matrix completes the velocity mapping operation from equation one.

After computing the real velocities (cm/s) at each pixel-coordinate inside the region of interest (aortic annulus), the mean blood flow velocity at each frame can be obtained. The application of equation three results in the calculation of blood volume that flows through the aortic annulus per frame. Plotting the blood volume per frame over a heart cycle yields the blood flow curve, whereas negative values correspond to regurgitant blood volume and positive values to forward blood volume, making possible the calculation of forward left ventricular stroke volume, regurgitant volume, total net volume and regurgitation fraction over the aortic annulus. Additional measurements include the aortic annulus major/minor diameters, perimeter and area, as described in **Figure 2**.

### Pixelwise velocity outlier correction — noise correction

One factor acknowledged to have an influence on error in phase-contrast MRI is random noise^51,52,53^. After applying velocity mapping on the dataset, we further refined the software performance after benchmarking multiple noise correction (pixel-intensity outlier removal) methods described here: first - computing the pixel-wise standard deviation over a heart cycle and thresholding the pixel-intensities to 0 if the standard-deviation was higher than the 99.95% percentile, with no further transformation otherwise; second - thresholding the pixel-intensities to 0 if the pixel-intensity over a heart cycle was lower than the 0.05% percentile, with no further transformation otherwise; third - setting the pixel-intensities to 0, if the pixel-intensity corresponded to > 200 cm/s or < −100 cm/s. The method achieving the highest Pearson correlation coefficient comparing calculated forward left ventricular stroke volume with our algorithm and Medviso Segment software (test set of 6,000 images from 200 UK Biobank individuals, described in the next section) was integrated into the pipeline (**Supplementary table 4** and **supplementary figures 1 - 3** show the software performance results).

### Benchmarking and quality control

Medviso’s open-source segmentation software^13^ was used for the post processing of phase contrast magnetic resonance imaging sequences to evaluate the aortic valve regurgitation fraction and forward volume of 200 randomly selected participants (not included for model segmentation training/validation/testing described in step 1) of the UK Biobank. The Medviso’s Segment software semi-automatically obtains these measurements using vessel-tracking algorithms, where one segmentation mask (which requires human intervention) is propagated to the remaining frames of the heart cycle. The most common approach of current commercially available scanner software tools relies on vessel tracking after careful manual segmentation of one frame, which is mostly done in the mid-systole, where the aortic annulus contours display the highest contrast. Due to the lack of a manually curated phase contrast magnetic resonance imaging dataset from the UK Biobank, we decided to design the ground-truth label of the test set based on the normally performed MRI analysis based on a human supervised vessel tracking technique with the open-source Medviso Segment, to closely mimic clinical practice.

This test set compilation was accomplished by a MRI level III trained physician (BG). The main performance metric includes the Pearson correlation coefficient (also averaging Pearson correlation coefficient from 5 complementary testing subsets, each with samples from 40 individuals) and the accompanying means and mean-differences obtained through Bland-Altman plots.

To ensure cross-platform compatibility, rapid setup, and to allow stringent control over versioning, we adopted a Docker-based strategy to containerize the program along with its dependencies. This method radically reduces the turnaround time for the software installation by incorporating all the program code and dependencies in one place. Additionally, using Docker, we could build specific optimized packages for multiple system architectures. The first one was built on the most widely used x86_64 architecture and comes in both GPU and CPU variants, with the latter being substantially smaller in size. There was no noticeable difference between the two versions in our internal tests using a Linux laptop with a 16-core CPU and an 8 GB NVIDIA RTX 2080 Super GPU, but this might radically alter in favor of the GPU version when numerous desktop/server level GPUs are accessible to the program. The other Docker container was built on aarch64 architecture, which natively supports the latest macOS® systems using Apple® silicon (M1/ M2) along with other ARM64 based systems. This software container, tested on an Apple MacBook Pro with M1 Max processor and 64GB of RAM, yielded almost identical processing speeds as our Linux test system. While the same was not true when we tried running our software container with x86_64 architecture in emulation mode, hence proving the benefits of building on native architecture. The software accepts a folder location carrying all DICOM ZIP files (one per sample) and creates a single tab-delimited list of results for all samples run in one batch. Additionally, a flow-over-aorta *vs* time graph is plotted for each sample.

DeepFlow processing on our test device took about 2 minutes and 30 seconds per phase contrast magnetic resonance imaging study. It is compatible with all major modern Linux, macOS, and Windows distributions. The software is freely available for non-commercial use (open-source CC-BY-NC license) at https://github.com/Urban90/deepFlow.

### Outcomes analysis

We focused our outcomes analysis on aortic valve regurgitation fraction, as we sought to further validate DeepFlow’s phenotyping. In order to ensure that no participant had more than one MRI measurement included in the study, only the first phase contrast magnetic resonance imaging visit was included. The means and standard deviations were used to infer the prevalence of aortic valve regurgitation in the UK Biobank. Student’s T-tests for independent variables were applied to compare the means of aortic valve regurgitation distribution among age groups and between sex categories. To infer progression rates per year of life, multivariate linear regression models to predict aortic valve regurgitation fraction were constructed. The first used sex and age as covariates, the second model added ascending aortic aneurysm (obtained after binarizing the variable of maximal aorta diameter computed by the developed software, using a cut-off of 50 mm or higher), the third added presence of hypertension (instead of ascending aortic aneurysm). Also, a multivariate linear regression model including age at MRI, sex, presence of hypertension and presence of aneurysm to predict aortic valve regurgitation fraction was computed (**Supplementary discussion SD1**). To assess the distribution of common cardiovascular comorbidities across the aortic valve regurgitation spectrum categories (mild: < 22%; moderate >= 22% and < 33%; severe >= 33%), Chi-Square tests were applied. Considering these statistical analyses, a two-sided level of significance of 0.05 was adopted.

Comorbidities surrogates were defined as a binary categorical variable, using UK Biobank’s International Classification of Diseases 10th Revision (ICD-10) codes for intra-hospital diagnoses. These common cardiovascular comorbidities included coronary artery disease, valvular heart disease, non-ischemic cardiomyopathies, inflammatory heart diseases, atrial fibrillation, diabetes mellitus type 2, hypertension, chronic obstructive pulmonary disease and peripheral artery disease. Chi-Square test results, adopted ICD-10-codes and distribution summary are compiled in **Table 1**. The distribution of congenital aortic valve malformations (per ICD-10 codes), ascending aorta aneurysm and rheumatic aortic valve disease (as per ICD-10 code) across the three groups of aortic valve regurgitation severity are outlined in **Table 2**. Also, the history of procedures were analyzed, using the operative procedure codes (OPCS-4). Main procedures included coronary revascularizations (surgical and interventional), heart valve repair or replacement (surgical or interventional), heart transplantation, left ventricular assist devices, permanent pacemaker implantation and the presence of an implantable cardioverter-defibrillator. Only the ICD-10 codes or OPCS-4 codes dated before MRI was performed were included. The distribution of procedures history and Chi-Square test results are illustrated in **Table 3**.

**Table 2.** Etiologic comorbidities across the aortic valve regurgitation fraction severity.

| **N = 33590** | **AR fraction** |  |  |  |
| --- | --- | --- | --- | --- |
| **Diagnosis (ICD10-code)** | **Mild**<br>**(< 22%)** | **Moderate**<br>**(>= 22% & < 33%)** | **Severe**<br>**(>= 33%)** | **p-value** |
| **Rheumatic valve disease**<br>**(I.06)** | 1 | 0 | 1 | 1e<sup>-11</sup> *** |
| **Congenital aortic valve disease**<br>**(Q.23)** | 4 | 0 | 0 | 0.99 |
| **Aneurysm of the ascending aorta** | 165 | 16 | 30 | $1.8e^{-113}$ *** |

ICD10-codes definitions are considered as surrogate markers of the comorbidities. \*\* indicates p-value < 0.01, \*\*\* indicates p-value < 0.0001.

**Table 3.**
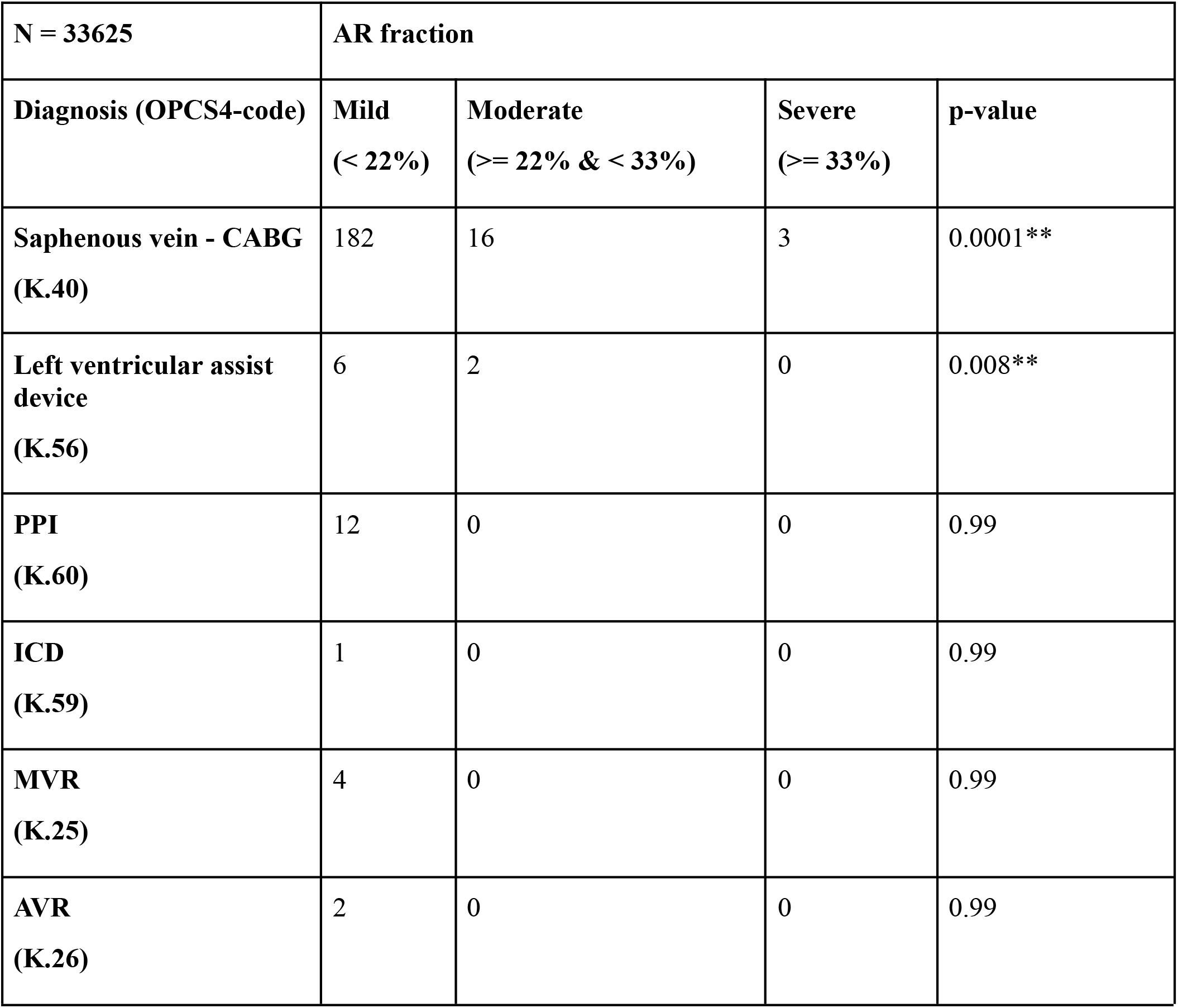
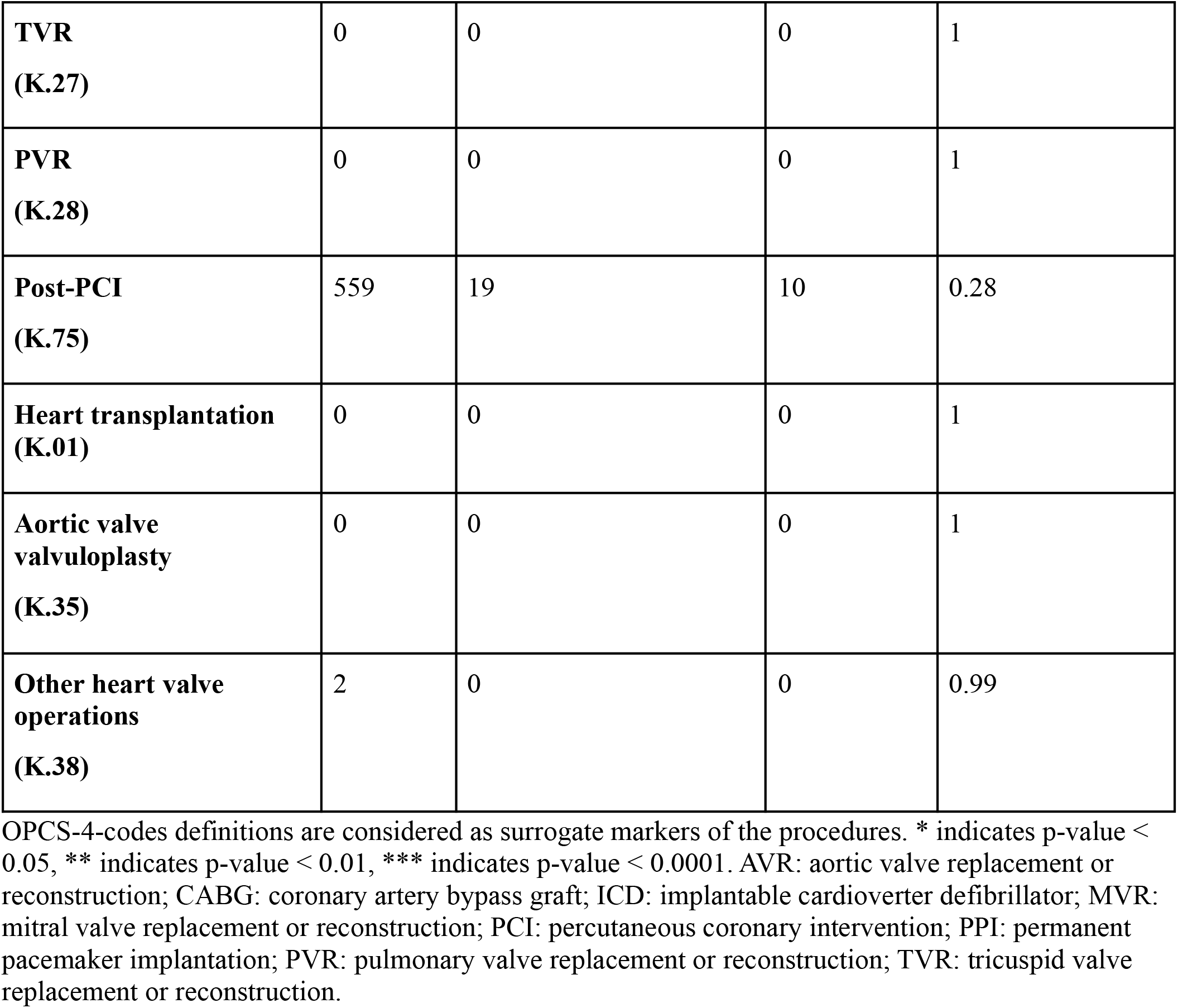
Baseline history of cardiovascular procedures across the aortic valve regurgitation fraction severity.

| **N = 33625** | **AR fraction** |  |  |  |
| --- | --- | --- | --- | --- |
| **Diagnosis (OPCS4-code)** | **Mild**<br>**(< 22%)** | **Moderate**<br>**(≥ 22% & < 33%)** | **Severe**<br>**(≥ 33%)** | **p-value** |
| **Saphenous vein - CABG**<br>**(K.40)** | 182 | 16 | 3 | 0.0001** |
| **Left ventricular assist device**<br>**(K.56)** | 6 | 2 | 0 | 0.008** |
| **PPI**<br>**(K.60)** | 12 | 0 | 0 | 0.99 |
| **ICD**<br>**(K.59)** | 1 | 0 | 0 | 0.99 |
| **MVR**<br>**(K.25)** | 4 | 0 | 0 | 0.99 |
| **AVR**<br>**(K.26)** | 2 | 0 | 0 | 0.99 |
| **TVR**<br>**(K.27)** | 0 | 0 | 0 | 1 |
| **PVR**<br>**(K.28)** | 0 | 0 | 0 | 1 |
| **Post-PCI**<br>**(K.75)** | 559 | 19 | 10 | 0.28 |
| **Heart transplantation**<br>**(K.01)** | 0 | 0 | 0 | 1 |
| **Aortic valve**<br>**valvuloplasty**<br>**(K.35)** | 0 | 0 | 0 | 1 |
| **Other heart valve**<br>**operations**<br>**(K.38)** | 2 | 0 | 0 | 0.99 |

OPCS-4-codes definitions are considered as surrogate markers of the procedures. \* indicates p-value < 0.05, \*\* indicates p-value < 0.01, \*\*\* indicates p-value < 0.0001. AVR: aortic valve replacement or reconstruction; CABG: coronary artery bypass graft; ICD: implantable cardioverter defibrillator; MVR: mitral valve replacement or reconstruction; PCI: percutaneous coronary intervention; PPI: permanent pacemaker implantation; PVR: pulmonary valve replacement or reconstruction; TVR: tricuspid valve replacement or reconstruction.

**Table 4.** Mendelian randomization analysis

|  |  |
| --- | --- |
| **Two-Sample Mendelian**<br>**Randomization Analysis**<br>**(non overlapping subsets)** | **Exposure: aortic annulus area (N= 17772)**<br>**Outcome: aortic valve regurgitation fraction (N= 18988)** |
| MR Egger | Beta = 0.23 ± 0.08, P-value = 5.6e<sup>-3</sup> |
| Weighted median | Beta = $0.19 \pm 0.04$ P-value = $1.4e^{-7}$ |
| Inverse variance weighted | Beta = $0.17 \pm 0.03$ P-value = $5.6e^{-10}$ |

Mendelian Randomization analysis establishes a causal relationship between BSA-indexed aortic annulus area (exposure) and aortic valve regurgitation fraction (outcome) using two sample Mendelian randomization techniques on non-overlapping samples for robustness of results. A p-value is considered significant if $< 0.05$ . N indicates sample size. Beta indicates the effect size ± standard error.

### Time-to-event analysis

A time-to-event analysis of several clinical endpoints was performed. Clinical endpoints included all-cause mortality, ischemic stroke, coronary artery bypass graft and aortic valve replacement. The first two outcomes were obtained based on a composite source of data from an interview with a nurse at the visit to assessment centers (self-reported) and linked electronic health records including hospital inpatient episode data. Hospital inpatient episode data was collected at the Assessment Centre in-patient Health Episode Statistics in combination with data on cause of death from the National Health Service Information Centre. The remaining outcome endpoints were retrieved by the date the respective ICD-10-code was documented in the intra-hospital electronic medical record of the participant. Only ICD-10-codes recorded after the date of the MRI visit were taken into account. All participants without the following cardiovascular comorbidities (using the ICD-10 codes): I.10, I.25, I.34 to I.37, I.40 to I.43, I.48 and I.73; and/or procedures (using the OPCS-4 codes): K.01, K.25 to K.28, K.30, K.35, K.38, K.40, K.41, K.56, K.59, K.60 and K.75 were excluded from the subsequent analysis. Tables **1** to **3** show the ICD-10 codes/OPCS-4 codes corresponding clinical conditions. All missing values were excluded from the analysis and a Z-score standardization of the remaining dataset was performed. Also, aortic valve regurgitation was adjusted to age and sex (excluding the residuals computed after fitting a linear regression model to these covariates). Aortic valve regurgitation fraction was transformed into a categorical variable (mild: < 22%; moderate >= 22% and < 33%; severe >= 33%). Ischemic stroke and coronary artery bypass graft related results are presented in the **supplementary material S2**. Excluding the time-to-event analysis of the outcome of new aortic valve replacement at follow-up as the dependent variable, the event of aortic valve replacement was right-censored. Then, Kaplan-Meyer curves and log-rank tests were undertaken to analyze the possible differences in clinical outcomes rates across aortic valve regurgitation severity groups. For these statistical analyses, a two-sided level of significance of 0.05 was adopted.

### Genome-wide association studies

We studied the genotype-phenotype associations in the cardiac flow related variables extracted from raw phase-contrast magnetic resonance imaging after deploying DeepFlow to the available UK Biobank cardiac imaging database. Also, to compute the genetic correlation matrix including the remaining left and right ventricular morphological and functional traits, a published deep-learning based model^4^ was used. Mitral valve regurgitation volume was calculated using the forward left ventricular stroke volume (*AoPC*) output from the developed software (phase contrast magnetic resonance imaging based) and the total left ventricular stroke volume (*LVSV*) from the Bai et al.’s model (CINE sequence based), after equation seven:

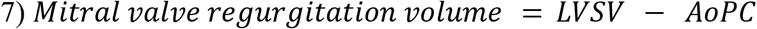

Other MRI-based mitral valve regurgitation quantification methods include (equations 8 and 9):

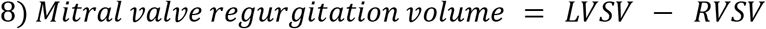

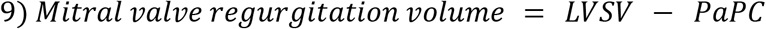

Although prognostic and diagnostic outcome data regarding mitral valve regurgitation are mostly based on the method following equation 7 and is commonly preferred in the clinical setting, one limitation is the necessary combination of two acquisitions, subject to potential interscan variability^11^. Contrary to the methods from equations 8 and 9, mitral valve regurgitation quantification using equation 7 is independent from biases resulting from accompanying cardiac valvular pathology (which are an important source of error in method following equation 8) or from intracardiac shunts (e.g. an atrial septal defect causing left-right shunt; which leads to measuring errors while using equation 9).

Regarding the genetic data, participants were genotyped using custom arrays, Affymetrix UK BiLEVE Axiom array or Affymetrix UK Biobank Axiom array which shares 95% of marker content. The genetic data was centrally imputed into the Haplotype Reference Consortium panel and the UK10K+1000 Genomes panel^47^. Variant positions were identified using the GRCh37 human genome reference. The genotyping methods, arrays and quality-control procedures have been extensively described previously^54^. Only autosomes were included in the analysis.

Only participants with phase contrast magnetic resonance imaging sequences and genotyping through custom arrays were considered for analysis. Phenotype outliers defined as values more than three interquartile ranges above the first quartile or below the third quartile were winsorized. The residuals from the phenotypes’ variables were computed after linear regression models were fitted to the covariates (age at MRI, sex, the first 5 principal components and the genotyping array) and quantile normalization was applied. The following genetic quality control (QC) criteria were applied: filtering variants with a minor allele frequency less than 1%, filtering variants with a low minimal allele count (less than 20), excluding variants with missing call rates exceeding 1%, excluding variants failing a Hardy-Weinberg equilibrium exact test at 1\**e*^−6^. We performed a common variant genome wide association analysis of the traits using a mixed linear regression model through the SAIGE software^55^. To adjust for multiple testing, Bonferroni correction using the genome-wide significance threshold of p-value < 5 × 10^−8^ for the selection of independent variants that were significantly associated with the phenotypes was applied. Each phenotype underwent linkage disequilibrium (LD)-based clumping to generate independent SNVs using an LD cut-off of R^2^ < 0.1 within 500 kilobases window on variations exceeding the P-value threshold of 1* 10^−8^. Manhattan plots and QQ plots were generated using LocusZoom software^56^. SNV-heritability and genetic correlations between cardiac measurements were computed with LDSC software^20^.

Additionally, we carried out gene clustering using K-means (n = 3) and additional enrichment analysis for pathways and phenotypes using the STRING-DB server^25^ in order to comprehend the gene interactions of the major protein coding loci from Ao_F_Vmax and aortic annulus area traits. Additionally, STRING will run an automated pathway enrichment analysis of the gene clusters and identify any significantly associated pathways or phenotypes (using hypergeometric testing, against a statistical background of either the entire genome or a user-supplied background gene list). STRING performs these tests for a total of eleven functional pathway classification frameworks, including the Gene Ontology ^26^ and Monarch Human Phenotype Ontology (HPO)^27^. The STRING database seeks to incorporate all known and anticipated relationships between genes/proteins, including both functional and physical interactions. STRING gathers and scores data from a variety of sources to accomplish this, including automated text mining of the scientific literature, databases of interaction experiments and annotated complexes/pathways, computational interaction predictions from co-expression and from conserved genomic context, and systematic transfers of interaction evidence from one organism to another^57^.

### Mendelian randomization

We evaluated the bidirectional causal effects between aortic annulus area and aortic valve regurgitation fraction with the Bayesian-based Mendelian randomization (MR) method CAUSE^58^. CAUSE takes into account both correlated and uncorrelated pleiotropy, thereby mitigating the risk of horizontal pleiotropy bias. CAUSE permits overlapping GWAS datasets, and thus can be used both for single- and two-sample MR. Given a significant CAUSE model, we used the estimated difference in the expected log pointwise posterior density (delta ELPD, where the biggest negative difference supports the causal model) to rank variants based on their contribution to the causal estimate. As input instrumental variables for each MR analysis, we considered all SNVs with a P-value threshold of 5 * 10^−5^ or lower when examining the exposure genome-wide association results. We then applied LD-clumping using PLINK (default settings recommended by CAUSE: LD cut-off of R^2^ < 0.01 within 10000 kilobases window).

If CAUSE indicated a significant causal relationship (at p<0.05) for a given exposure-outcome pair, we carried the additional two-sample MR analysis explained below. We first randomly selected two non-overlapping subsets of the tested UK Biobank participants (e.g., ∼ 1:1 GWAS sample-size ratio between the traits aortic valve regurgitation and aortic annulus area). Using the TwoSample MR software^59^, summary data for exposure and outcomes were harmonized. When feasible, palindromic SNVs were deleted if the minor allele frequency was more than 0.42. We followed the suggested parameters for inferring forward strand alleles using allele frequency data. To assess for horizontal pleiotropy we used the built-in function of the TwoSample MR software to return the intercept values and respective p-values (non significant if p-value > 0.05) after applying the Egger-estimator. Finally, we used the inverse variance weighted random effects model, as well as the weighted median and the Egger-estimator to obtain causal effects and their significance.

In order to assess the MR assumption that genetic instruments are connected to the outcome only through the exposure, we also tested for reverse causation using MR-Steiger. Briefly, when reverse causation does not occur, the genetic mutation is more strongly related to the exposure. However, when reverse causation occurs, this premise is broken. We therefore also report the output of MR-Steiger directionality test estimates in the results section.

### Tissue enrichment analysis

In order to highlight biological pathways and sets of functionally related genes enriched by multiple GWAS signals; to identify tissues and cell types where prioritized genes are highly expressed we used DEPICT^30,31^, a tool that employs data from massive numbers of experiments measuring gene expression. For Linkage-Disequilibrium-based clumping by PLINK (v1.9), which precedes the DEPICT analysis, a *p* value threshold of 10^−8^ (except for aortic valve regurgitation fraction, whereas a p value threshold of 5*10^−5^ was established), a distance threshold of 500 kilobases and a Linkage-Disequilibrium threshold of 0.1 was set (default setting by DEPICT). We performed these analyses on the newly studied phenotypes with higher heritability **—** aortic annulus area, mean forward peak velocity at the aortic annulus, aortic valve regurgitation fraction.

