## Supplementary material for "Genetic architecture of cardiac dynamic flow volumes"

### Supplementary tables

**Table S1.** Hyperparameters of the batch normalization layers of DeepFlow's segmentation model

| | **Batch normalization** | | | |
|---|---|---|---|---|
| **Hyperparameters** | **gamma** | **beta** | **Moving mean** | **Moving variance** |
| **Layer 1** | 16 | 16 | 16 | 16 |
| **Layer 2** | 32 | 32 | 32 | 32 |
| **Layer 3** | 64 | 64 | 64 | 64 |
| **Layer 4** | 128 | 128 | 128 | 128 |
| **Layer 5** | 256 | 256 | 256 | 256 |
| **Layer 6** | 128 | 128 | 128 | 128 |
| **Layer 7** | 64 | 64 | 64 | 64 |
| **Layer 8** | 32 | 32 | 32 | 32 |
| **Layer 9** | 16 | 16 | 16 | 16 |

**Table S2.** Hyperparameters of the convolutional layers with a 3x3 kernel of DeepFlow's segmentation model

| | **2D Convolutional layer (kernel 3x3, stride = 2, padding = same)** | |
|---|---|---|
| **Hyperparameters** | Kernel | Bias |
| **Layer 1** | $3 \times 3 \times 1 \times 16$ | 16 |
| **Layer 2** | $3 \times 3 \times 16 \times 32$ | 32 |
| **Layer 3** | $3 \times 3 \times 32 \times 64$ | 64 |
| **Layer 4** | $3 \times 3 \times 64 \times 128$ | 128 |
| **Layer 5** | $3 \times 3 \times 128 \times 256$ | 256 |

| | | |
|---|---|---|
| **Layer 6** | $3 \times 3 \times 256 \times 128$ | 128 |
| **Layer 7** | $3 \times 3 \times 128 \times 64$ | 64 |
| **Layer 8** | $3 \times 3 \times 64 \times 32$ | 32 |
| **Layer 9** | $3 \times 3 \times 32 \times 16$ | 16 |

**Table S3.** Hyperparameters of the convolutional layers with a 1x1 kernel of DeepFlow's segmentation model

| | **2D Convolutional layer (kernel 1x1 , stride = 2, padding = same)** | |
|---|---|---|
| **Hyperparameters** | Kernel | Bias |
| **Layer 9** | $1 \times 1 \times 16 \times 1$ | 1 |

**Table S4.** DeepFlow model performance applying different Noise (pixelwise velocity outlier) minimization methods.

| **DeepFlow postprocessing**<br>**Noise (pixelwise velocity outlier) minimization methods** | **Forward LV stroke volume (Pearson correlation ± standard deviation)** | **Aortic valve regurgitation fraction (Pearson correlation ± standard deviation)** |
|---|---|---|
| Setting pixel-intensities to 0 if the standard-deviation over the heart cycle was higher than the 99.95% percentile at that pixel-coordinate | 0.93 ± 0.01*** | 0.91 ± 0.03*** |
| Setting pixel-intensities to 0 if the pixel-intensity over a heart cycle was lower than the 0.05% percentile at that pixel-coordinate | 0.96 ± 0.02*** | 0.90 ± 0.03*** |

| | | |
|---|---|---|
| Setting the pixel-intensities to 0, if the pixel-intensity corresponded to > 200 cm/s or < -100 cm/s | 0.95 ± 0.02*** | 0.88 ± 0.02*** |

Testset included 6000 images from 200 individuals (30 images per individual) of the UK Biobank, compared after manual curation using the Medviso software. LV indicates left ventricle. The average Pearson correlation coefficient from 5 complementary testing subsets, each with samples from 40 individuals, was computed. ***indicates that the P-value of the calculated Pearson correlation for each subset of 40 individuals was lower than 0.001.

**Table S5.** Linkage-Disequilibrium Score regression intercepts and respective standard deviations of the GWAS for different cardiac dynamic flow and volume phenotypes.

| **Cardiac dynamic flow and volumes phenotypes** | **LD regression score intercept** | **Standard deviation** |
|---|---|---|
| **Aortic valve regurgitation fraction** | 1.0218 | 0.0102 |
| **Aortic valve regurgitation volume** | 1.0164 | 0.0095 |
| **Total left ventricular stroke volume** | 1.0062 | 0.0105 |
| **Forward left ventricular stroke volume** | 1.0054 | 0.0104 |
| **Net left ventricular stroke volume** | 1.0145 | 0.0103 |
| **Forward peak velocity at aortic annulus** | 1.0254 | 0.0125 |
| **Regurgitant peak velocity at aortic annulus** | 1.0118 | 0.0099 |
| **Aortic annulus area** | 1.0146 | 0.0113 |

**Table S6.** Genomic Loci Identified for BSA-indexed aortic annulus area

| SNV | Chromosome | Position | Gene loci | P-Value |
|---|---|---|---|---|
| rs11768878 | chr7 | 73431169 | *ELN* | 1.8E-53 |
| rs7336555 | chr13 | 22863272 | *MTND3P1* | 1.22E-33 |
| rs6895341 | chr5 | 95617095 | *CTD-2337A12.1* | 6.33E-30 |
| rs17470137 | chr5 | 122531347 | *PRDM6* | 1.15E-23 |
| rs627634 | chr15 | 48879291 | *FBN1* | 5.62E-23 |
| rs9848754 | chr3 | 41753647 | *ULK4* | 7.03E-20 |
| rs77870048 | chr16 | 69965021 | *WWP2* | 5.49E-19 |
| rs4078169 | chr5 | 95454983 | *RP11-254I22.1* | 4.28E-18 |
| rs216193 | chr17 | 2203453 | *SMG6* | 9.06E-18 |
| rs861202 | chr12 | 21994955 | *ABCC9* | 2.84E-17 |
| rs861202 | chr12 | 21994955 | *RP11-729I10.2* | 2.84E-17 |
| rs72931748 | chr11 | 69825414 | *RP11-626H12.1* | 6.56E-17 |
| rs10202552 | chr2 | 19709049 | *AC010096.2* | 1.07E-16 |
| rs7973748 | chr12 | 20164705 | *RP11-405A12.2* | 1.35E-16 |
| rs1441358 | chr15 | 71612514 | *THSD4* | 4.47E-15 |
| rs2070699 | chr6 | 12292772 | *EDN1* | 7.73E-15 |
| rs34925856 | chr10 | 95927267 | *PLCE1* | 2.05E-14 |
| rs4821942 | chr22 | 40718100 | *TNRC6B* | 3.9E-14 |
| rs4264431 | chr17 | 12181294 | *RP11-471L13.2* | 4.94E-14 |
| rs12771055 | chr10 | 96116420 | *NOC3L* | 5.5E-14 |
| rs2276068 | chr11 | 70007484 | *ANO1* | 1.9E-13 |
| rs140270414 | chr12 | 62761516 | *USP15* | 3.06E-13 |

| | | | | |
|---|---|---|---|---|
| rs77097530 | chr5 | 122609853 | *CTD-2347I17.1* | 4.15E-13 |
| rs72800819 | chr5 | 122422449 | *AC106786.1* | 6.93E-13 |
| rs13211322 | chr6 | 152398050 | *ESR1* | 1E-12 |
| rs62049035 | chr16 | 70491655 | *FUK* | 1.4E-12 |
| rs2760748 | chr17 | 2001604 | *RP11-667K14.5* | 1.88E-12 |
| rs59317921 | chr11 | 130270855 | *ADAMTS8* | 6.02E-12 |
| rs1979974 | chr4 | 146800815 | *ZNF827* | 7.64E-12 |
| rs2812228 | chr13 | 50773790 | *DLEU1* | 1.84E-11 |
| rs79013608 | chr12 | 20230940 | *RP11-664H17.1* | 3.34E-11 |
| rs3848233 | chr16 | 88992338 | *CBFA2T3* | 4.73E-11 |
| rs1779211 | chr10 | 18514120 | *CACNB2* | 8.2E-11 |
| rs6581958 | chr12 | 71101532 | *PTPRR* | 1.31E-10 |
| rs57675369 | chr5 | 95162419 | *RHOBTB3* | 2.34E-10 |
| rs13266971 | chr8 | 124584867 | *RP11-174I12.2* | 4.31E-10 |
| rs12188202 | chr5 | 81825115 | *CTD-2015A6.1* | 4.95E-10 |
| rs6806 | chr17 | 2284005 | *SGSM2* | 6.4E-10 |
| rs13098199 | chr3 | 58119882 | *FLNB* | 6.86E-10 |
| rs13002621 | chr2 | 164915279 | *AC092684.1* | 0.00000000198 |
| rs116458863 | chr5 | 81731050 | *ATP6AP1L* | 0.00000000209 |
| rs294601 | chr3 | 14832407 | *RP11-95M5.1* | 0.00000000284 |
| rs55834964 | chr15 | 79049766 | *ADAMTS7* | 0.00000000449 |
| rs11205303 | chr1 | 149906413 | *MTMR11* | 0.00000000463 |
| rs7820261 | chr8 | 108292355 | *ANGPT1* | 0.00000000499 |
| rs146040837 | chr8 | 75780736 | *RP11-758M4.4* | 0.00000000636 |
| rs698083 | chr3 | 186997742 | *MASP1* | 0.0000000751 |

| | | | | |
|---|---|---|---|---|
| rs72790196 | chr16 | 83005337 | *CDH13* | 0.0000000104 |
| rs72790196 | chr16 | 83005337 | *CTD-3253I12.1* | 0.0000000104 |
| rs11761337 | chr7 | 73495550 | *LIMK1* | 0.0000000106 |
| rs55914222 | chr3 | 128202943 | *GATA2* | 0.0000000125 |
| rs6550446 | chr3 | 37066373 | *MLH1* | 0.0000000167 |
| rs965821 | chr5 | 122137708 | *SNX2* | 0.0000000195 |
| rs55855614 | chr20 | 10604466 | *SLX4IP* | 0.0000000211 |
| rs5874371 | chr6 | 12578095 | *RP11-125M16.1* | 0.0000000221 |
| rs248337 | chr5 | 95208902 | *AC008592.5* | 0.0000000313 |

**Table S7.** Genomic Loci Identified for peak forward velocity at the aortic annulus

| SNV | Chromosome | Position | Gene loci | P |
|---|---|---|---|---|
| rs6460068 | chr7 | 73428671 | *ELN* | 5E-44 |
| rs7336555 | chr13 | 22863272 | *MTND3P1* | 3.62E-29 |
| rs1532292 | chr17 | 2097483 | *SMG6* | 3.85E-21 |
| rs4264961 | chr5 | 95617018 | *CTD-2337A12.1* | 2.49E-20 |
| rs1649371 | chr2 | 19710745 | *AC010096.2* | 8.55E-19 |
| rs2544450 | chr12 | 21994280 | *ABCC9* | 2.52E-18 |
| rs2544450 | chr12 | 21994280 | *RP11-729I10.2* | 2.52E-18 |
| rs111412755 | chr11 | 69819139 | *RP11-626H12.1* | 1.64E-17 |
| rs2764339 | chr10 | 95918544 | *PLCE1* | 1.17E-14 |
| rs2070699 | chr6 | 12292772 | *EDN1* | 6.43E-14 |
| rs11045044 | chr12 | 20257908 | *RP11-664H17.1* | 1.24E-13 |
| rs17470137 | chr5 | 122531347 | *PRDM6* | 1.65E-13 |
| rs2276068 | chr11 | 70007484 | *ANO1* | 3.99E-13 |

| | | | | |
|---|---|---|---|---|
| rs3811942 | chr5 | 95728440 | *PCSK1* | 2.43E-12 |
| rs77870048 | chr16 | 69965021 | *WWP2* | 2.56E-12 |
| rs2466790 | chr15 | 48899399 | *FBN1* | 6.75E-12 |
| rs1979974 | chr4 | 146800815 | *ZNF827* | 6.94E-12 |
| rs2760748 | chr17 | 2001604 | *RP11-667K14.5* | 6.54E-11 |
| rs5874371 | chr6 | 12578095 | *RP11-125M16.1* | 6.78E-11 |
| rs2066637 | chr13 | 50766446 | *DLEU1* | 7.68E-11 |
| rs34734251 | chr17 | 12180963 | *RP11-471L13.2* | 8.42E-11 |
| rs3848233 | chr16 | 88992338 | *CBFA2T3* | 9.89E-10 |
| rs9848754 | chr3 | 41753647 | *ULK4* | 0.00000000101 |
| rs2078339 | chr12 | 20161168 | *RP11-405A12.2* | 0.00000000185 |
| rs77097530 | chr5 | 122609853 | *CTD-2347I17.1* | 0.00000000236 |
| rs9479214 | chr6 | 152402326 | *ESR1* | 0.00000000337 |
| rs9876116 | chr3 | 37083740 | *MLH1* | 0.00000000341 |
| rs137963273 | chr11 | 69878643 | *RP11-626H12.2* | 0.00000000354 |
| rs6806 | chr17 | 2284005 | *SGSM2* | 0.00000000642 |
| rs71482305 | chr10 | 96119130 | *NOC3L* | 0.00000000764 |
| rs13266971 | chr8 | 124584867 | *RP11-174I12.2* | 0.0000000128 |
| rs6581958 | chr12 | 71101532 | *PTPRR* | 0.0000000168 |
| rs6077843 | chr20 | 10578542 | *SLX4IP* | 0.0000000181 |
| rs17323619 | chr22 | 40538717 | *TNRC6B* | 0.000000023 |
| rs76027228 | chr7 | 73422593 | *RP11-731K22.1* | 0.0000000237 |
| rs9311153 | chr3 | 37167504 | *LRRFIP2* | 0.000000033 |
| rs34336464 | chr3 | 58127827 | *FLNB* | 0.0000000341 |
| rs11855326 | chr15 | 71610835 | *THSD4* | 0.0000000398 |

| | | | | |
|---|---|---|---|---|
| rs7625133 | chr3 | 187010381 | *MASP1* | 0.0000000447 |

**Table S8.** Genomic Loci Identified for BSA-indexed total left ventricular stroke volume

| **SNP** | **Chromosome** | **Position** | **Gene loci** | **P-Value** |
|---|---|---|---|---|
| rs937277 | chr6 | 118673075 | *SLC35F1* | 0.00000000102 |
| rs11065987 | chr12 | 112072424 | *BRAP* | 0.0000000025 |
| rs4246224 | chr12 | 24784139 | *RP11-615I16.1* | 0.0000000193 |
| rs17630235 | chr12 | 112591686 | *TRAFD1* | 0.0000000207 |
| rs12889267 | chr14 | 21542766 | *ARHGEF40* | 0.0000000249 |
| rs72647864 | chr2 | 179647824 | *TTN* | 0.0000000425 |
| rs1994135 | chr12 | 33682405 | *RNU6-400P* | 0.0000000439 |
| rs8176685 | chr9 | 136138766 | *ABO* | 0.000000044 |

**Table S9.** Genomic Loci Identified for aortic valve regurgitation volume

| **SNP** | **Chromosome** | **Position** | **Gene loci** | **P-Value** |
|---|---|---|---|---|
| rs77097530 | chr5 | 122609853 | *AC010493.1* | 3.28e-08 |
| rs4719607 | chr7 | 2659951 | *IQCE* | 4.58e-08 |

**Table S10. Genomic Loci Identified for aortic valve regurgitation fraction**

| **SNP** | **Chromosome** | **Position** | **Gene loci** | **P-Value** |
|---|---|---|---|---|
| rs10176996 | chr2 | 19708692 | *LINC01808* | 1.28e-08 |

**Table S11.** Genomic Loci Identified for BSA-indexed forward left ventricular stroke volume

| **SNP** | **Chromosome** | **Position** | **Gene loci** | **P-Value** |
|---|---|---|---|---|
| rs4727470 | chr7 | 100509201 | *SLC12A9* | 6.22e-09 |

In supplementary tables S5 to S11, the locus name indicates the nearest annotated gene. Independent variables were adjusted to age, sex and the first 5 genetic principal components and genotyping array. To adjust for multiple testing, we applied Bonferroni correction using the genome-wide significance threshold of $P < 5 \times 10^{-8}$ for selection of independent SNVs that were considered significantly associated with the traits.

### Supplementary figures

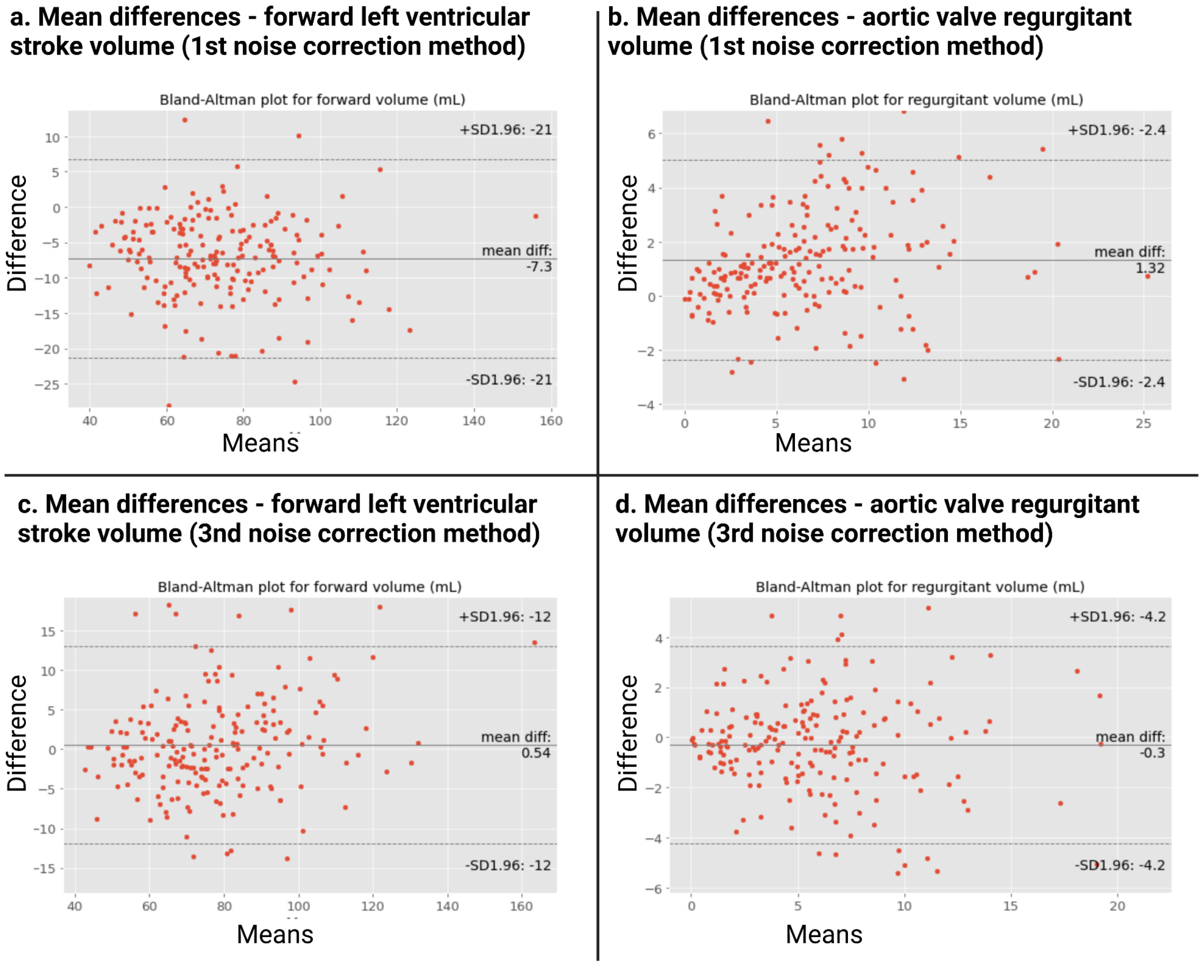


**Supplementary figure S1.** Bland-Altman plots for the analysis of the mean-differences in the outputs of forward left ventricular stroke volume (a) and (c), and aortic valve regurgitant volume (b) and (d) using the first and third noise minimization methods vs. the output using the Medvison Segment software, respectively. Mean diff indicates mean difference. SD indicates the Z-Score at 1.96, corresponding to a 95% confidence interval.

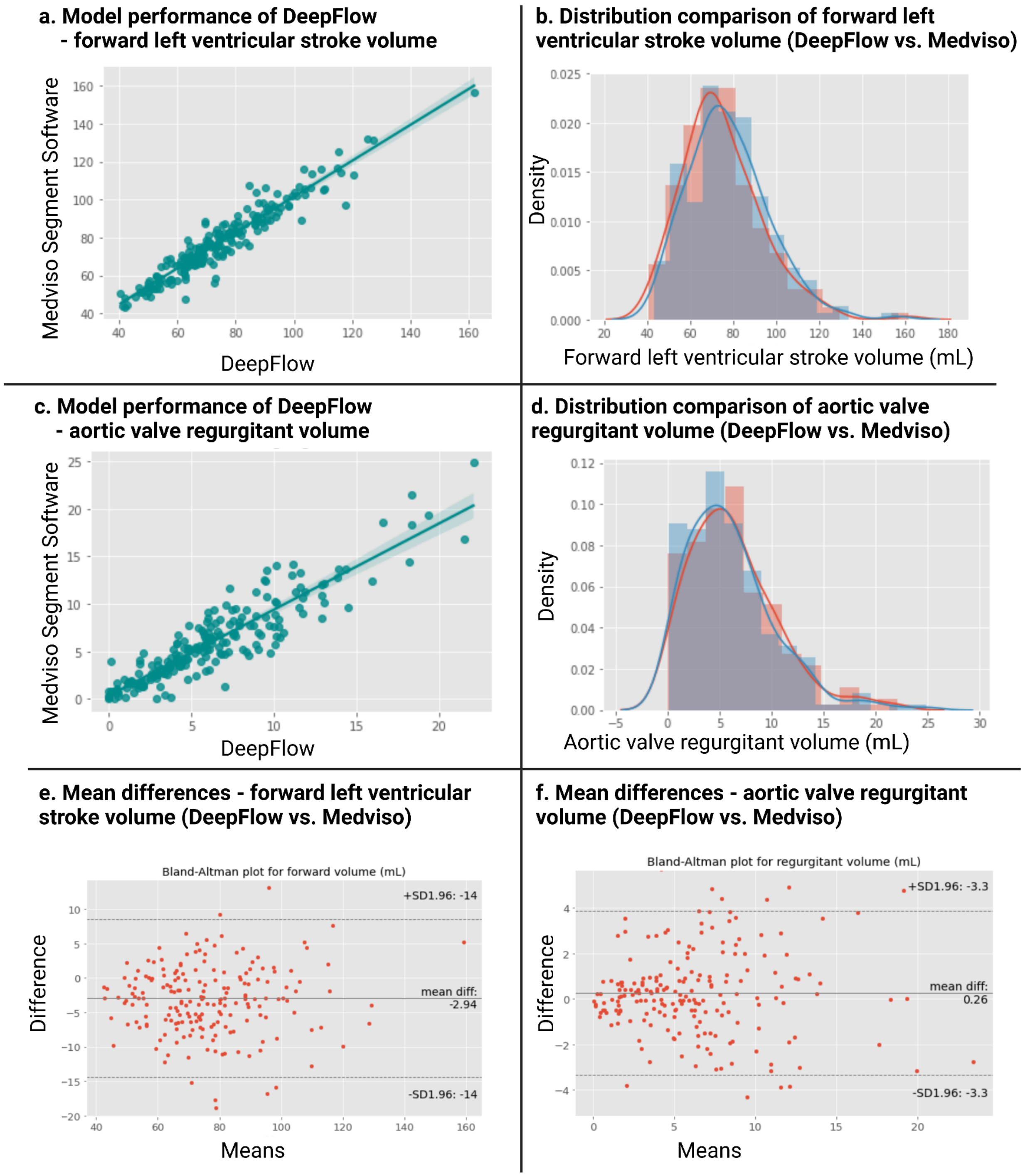
a. Model performance of DeepFlow - forward left ventricular stroke volume
Medviso Segment Software
DeepFlow
b. Distribution comparison of forward left ventricular stroke volume (DeepFlow vs. Medviso)
Density
Forward left ventricular stroke volume (mL)
c. Model performance of DeepFlow - aortic valve regurgitant volume
Medviso Segment Software
DeepFlow
d. Distribution comparison of aortic valve regurgitant volume (DeepFlow vs. Medviso)
Density
Aortic valve regurgitant volume (mL)
e. Mean differences - forward left ventricular stroke volume (DeepFlow vs. Medviso)
Bland-Altman plot for forward volume (mL)
+SD1.96: -14
mean diff: -2.94
-SD1.96: -14
Difference
Means
f. Mean differences - aortic valve regurgitant volume (DeepFlow vs. Medviso)
Bland-Altman plot for regurgitant volume (mL)
+SD1.96: -3.3
mean diff: 0.26
-SD1.96: -3.3
Difference
Means

**Supplementary figure S2.** DeepFlow performance using the best performing method of noise (velocity-encoding pixel outlier) optimization. The performance of DeepFlow software in predicting the aortic valve regurgitation fraction and the forward left ventricular stroke volume was most optimal when noise (pixelwise velocity outlier) correction was applied by reducing all pixel intensities < 0.05th percentile throughout the cardiac cycle to 0. Scatter plot (a) and density distribution histogram (b) of the correlation between the output of forward left ventricular stroke volume using the developed software (x-axis for scatter plot, red line for distribution histogram) vs. the output from the Medviso Segment software (y-axis for scatter plot, blue line for distribution histogram). Scatter plot (c) and density distribution histogram (d) of the correlation between the output of aortic valve regurgitant volume using the developed software vs. the output from the Medviso Segment software. Bland-Altman plots for the analysis of the mean-differences in the outputs of forward left ventricular stroke volume (e), and aortic valve regurgitant volume (f) using the developed software vs. the output from the Medviso Segment software. Mean diff indicates mean difference. SD indicates the Z-Score at 1.96, corresponding to a 95% confidence interval.

**a. Forward left ventricular stroke volume**

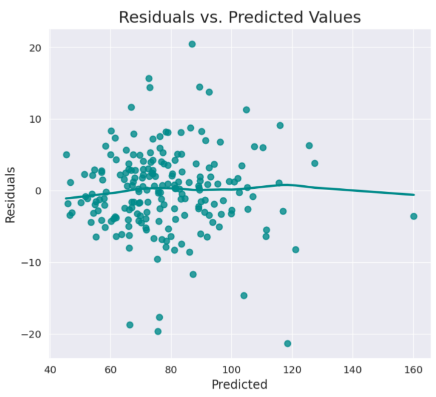


**b. Aortic valve regurgitantion volume**

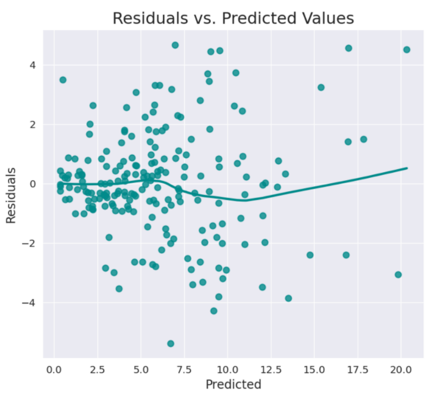


**Supplementary figure S3.** Distribution of the residuals compared with the predicted values of forward left ventricular stroke volume (a) and aortic valve regurgitation volume (b) by the developed software. In both cases, the sum of the residuals is close to zero ($4.48e^{-14}$ and $-1.25e^{-15}$, respectively) and follow close to an horizontal line, supporting the assumption of linearity between the variables (predictions from the developed software vs. predictions using the Medviso software).

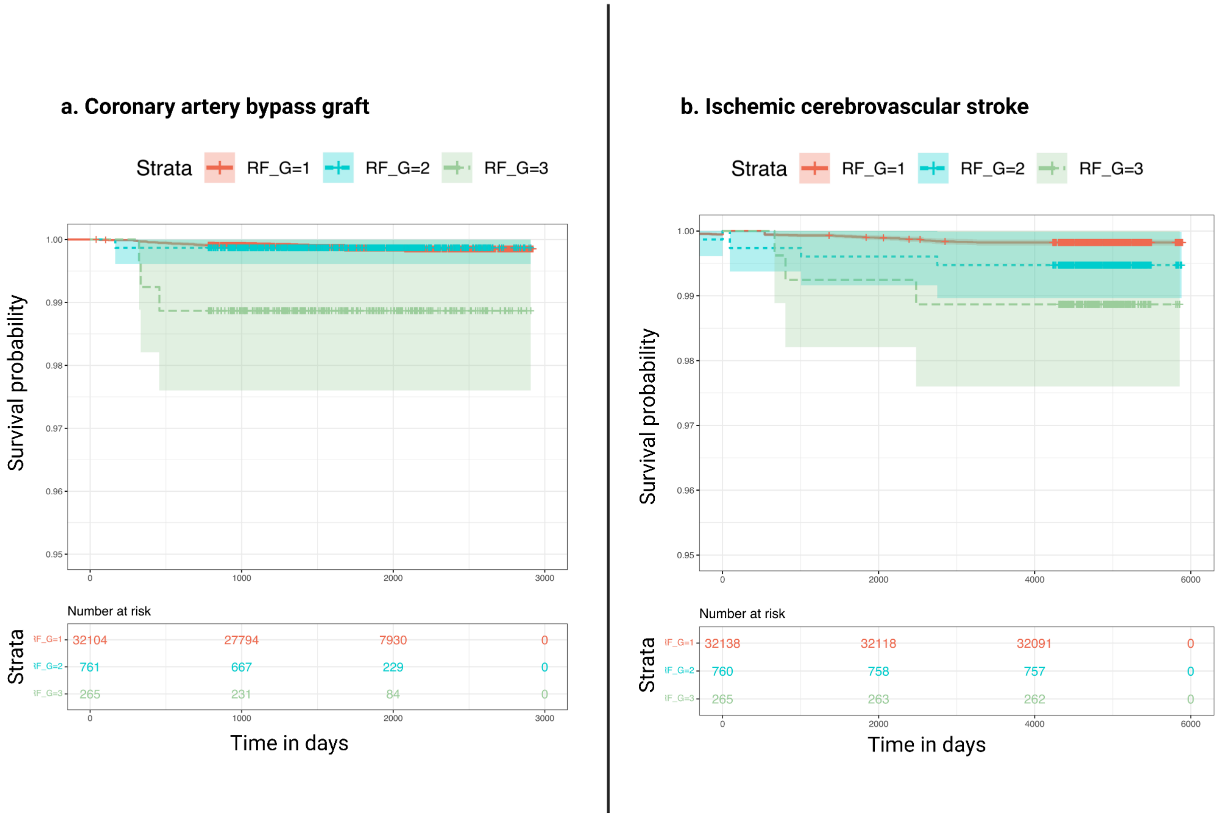


**Supplementary figure S4.** Time-to-event analysis of (a) new coronary artery bypass graft and (b) ischemic stroke in aortic valve regurgitation fraction. Common concomitant cardiovascular comorbidities were excluded (see Methods), and aortic valve regurgitation fraction was adjusted to sex and age at cardiac MRI. Log-rank test was performed to assess statistical significant differences between the incidence of outcomes. The percentiles of 99.2% for severe and 96.9% for moderate aortic valve regurgitation fraction were used to define the groups. Rates of coronary bypass grafts (mild vs. severe: $p < 0.005$ and moderate vs. severe: $p = 0.02$, log-rank test) and ischemic cerebrovascular stroke (mild vs. severe: $p = 0.006$, log-rank test) were enriched in patients with higher aortic regurgitation fractions. RF_G indicates aortic valve regurgitation fraction severity group: red: mild (1); blue: moderate (2); green: severe (3). Aortic valve replacement and coronary artery bypass graft surgeries were included as endpoints to analyze the discriminative power of DeepFlow's computed aortic valve regurgitation fraction.

**a. Aortic valve regurgitation fraction**

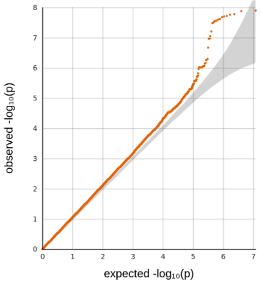


**GC lambda 0.5: 1.056**

**b. Aortic valve regurgitatant volume**

QQ Plot:

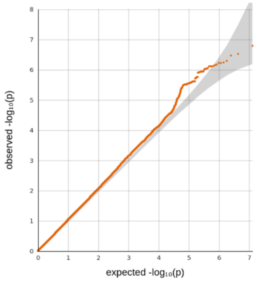


**GC lambda 0.5: 1.037**

**c. Mitral valve regurgitant volume**

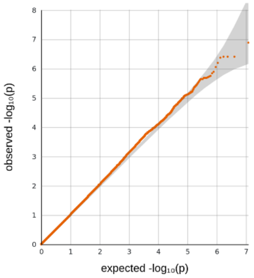


**GC lambda 0.5: 1.018**

**d. total LV stroke volume**

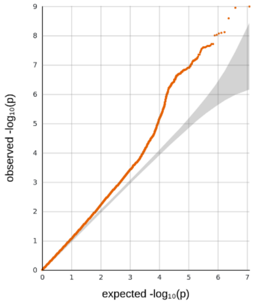


**GC lambda 0.5: 1.092**

**e. forward LV stroke volume**

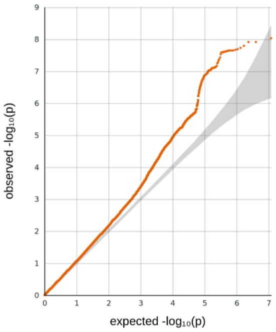


**GC lambda 0.5: 1.069**

**f. net LV stroke volume**

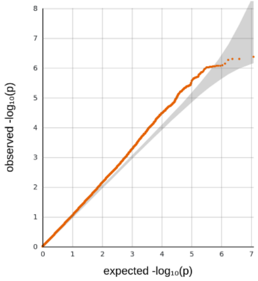


**GC lambda 0.5: 1.066**

**g. Aortic annulus area**

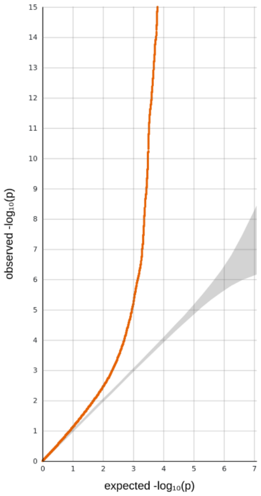


**GC lambda 0.5: 1.137**

**h. Forward peak velocity at aortic annulus**

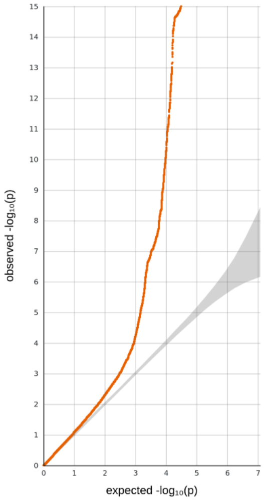


**GC lambda 0.5: 1.096**

**i. Regurgitant peak velocity at aortic annulus**

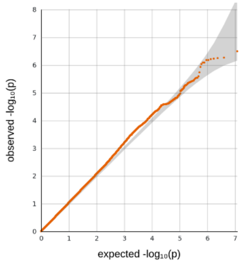


**GC lambda 0.5: 1.044**

**Supplementary figure S5.** QQ-plots of the phenotypes derived using DeepFlow software, as well as total left ventricular stroke volume and mitral valve regurgitation volume. All volumes and areas are BSA-indexed. The predicted distribution in -log10 is displayed on the X-axis (P-value). The observed distribution is shown on the Y-axis as -log10 (P-value). The red dots track the observed P-values in the performed GWAS, while the gray area depicts the expected P-values from a theoretical Chi-Square-distribution. LV indicates: left ventricular. The Genomic Control lambda (GC) calculated based on the 50th percentile (median). QQ-plots obtained with LocusZoom software.

**a. Cardiac phenotypes derived from DeepFlow**

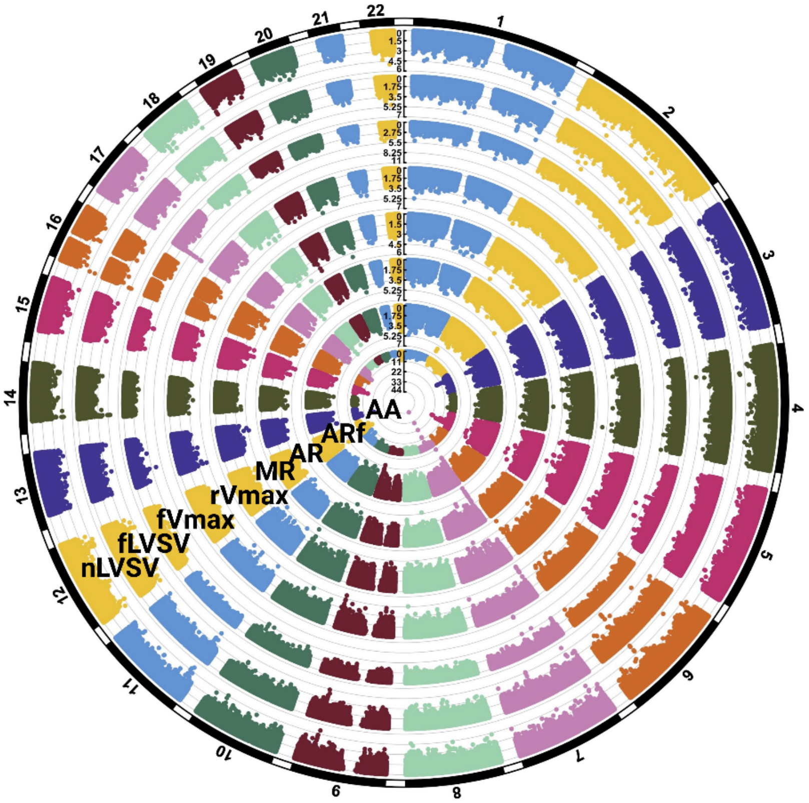


**b. Cardiac phenotypes derived from Bai *et al.* (CINE cardiac MRI)**

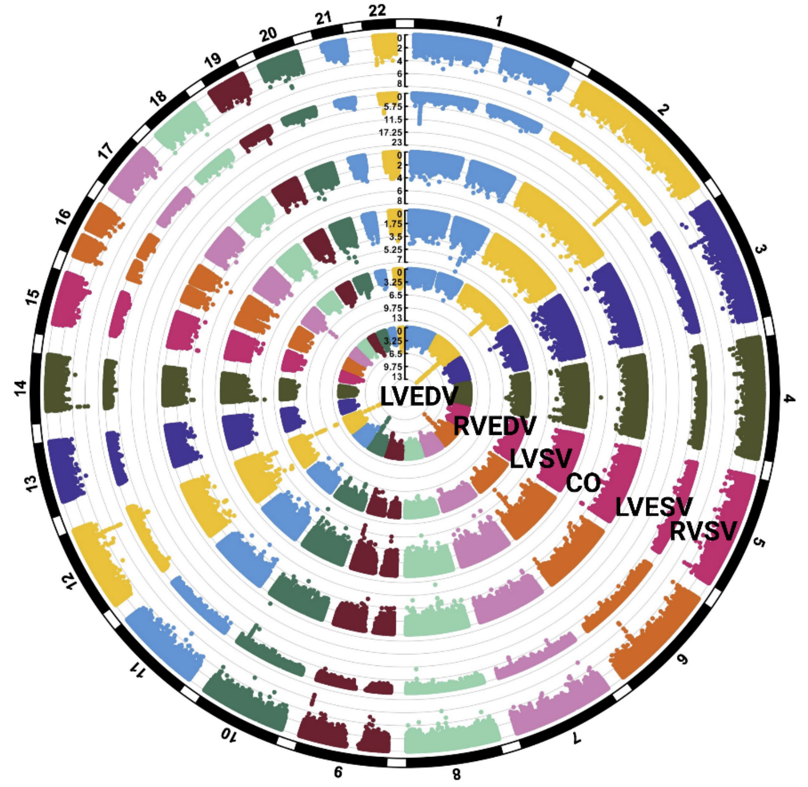

**Supplementary figure S6.** Circular Manhattan plot displaying the overview of GWAS results. In a GWAS results of the (a) phenotypes obtained from DeepFlow alone and mitral valve regurgitation volume are displayed. In (b) is a circular Manhattan plot of the GWAS results from phenotypes regarding LV and RV morphological and functional parameters obtained exclusively from Bai *et al.* model[4]. AA indicates: BSA indexed aortic annulus area; AR: aortic valve regurgitant volume; ARf: aortic valve regurgitation fraction; fVmax: aortic forward peak velocity; fLVSV: forward left ventricular stroke volume; MR: mitral valve regurgitant volume; nLVSV: net left ventricular stroke volume; rVmax: aortic retrograde peak velocity. CO: cardiac output from left ventricle; LVEDV: left ventricular end diastolic volume; LVESV: left ventricular end systolic volume; LVSV: total left ventricular stroke volume; RVEDV: right ventricular end diastolic volume; RVSV: right ventricular stroke volume. All volumes and areas are BSA indexed.

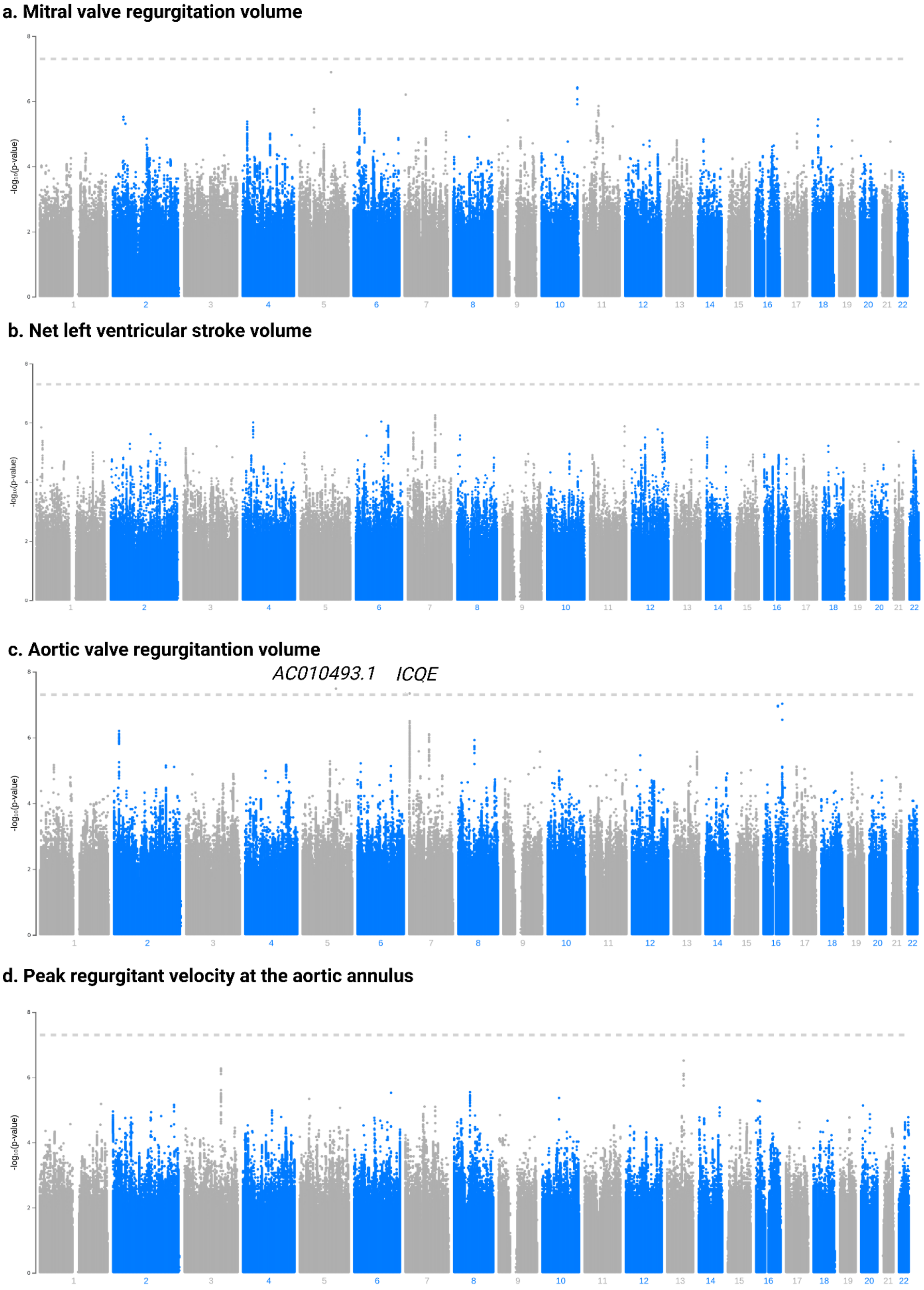
a. Mitral valve regurgitation volume
b. Net left ventricular stroke volume
c. Aortic valve regurgitantion volume
AC010493.1
ICQE
d. Peak regurgitant velocity at the aortic annulus
-log10(p-value)
1 2 3 4 5 6 7 8 9 10 11 12 13 14 15 16 17 18 19 20 21 22

**Supplementary figure S7.** Manhattan plots displaying the overview of GWAS results of the (a) mitral valve regurgitation volume, (b) net left ventricular stroke volume, (c) aortic valve regurgitation volume and (d) peak regurgitant velocity at the aortic annulus. Independent variables were adjusted to age, sex and the first 5 genetic principal components and genotyping array. To adjust for multiple testing, we applied Bonferroni correction using the genome-wide significance threshold of $P < 5 \times 10^{-8}$ for selection of independent SNVs that were considered significantly associated with the traits. All volumes and areas are BSA-indexed.

#### a. Aortic annulus area

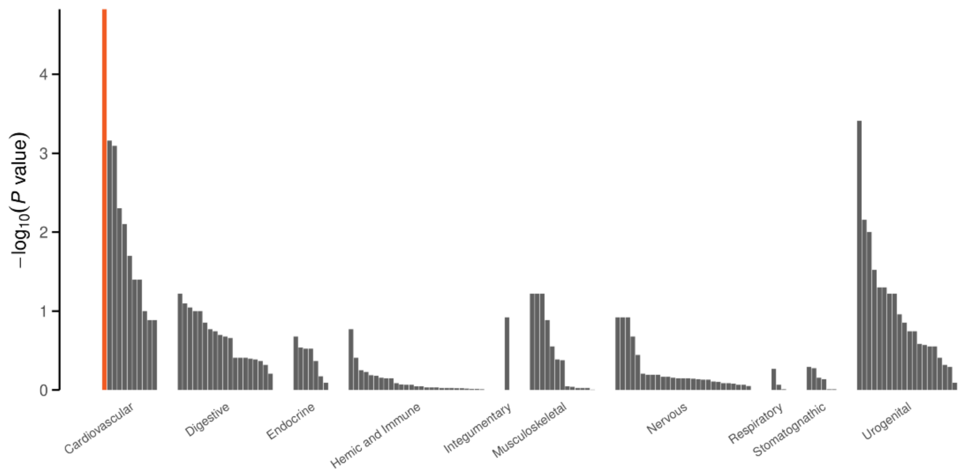


#### b. Forward peak velocity at aortic annulus

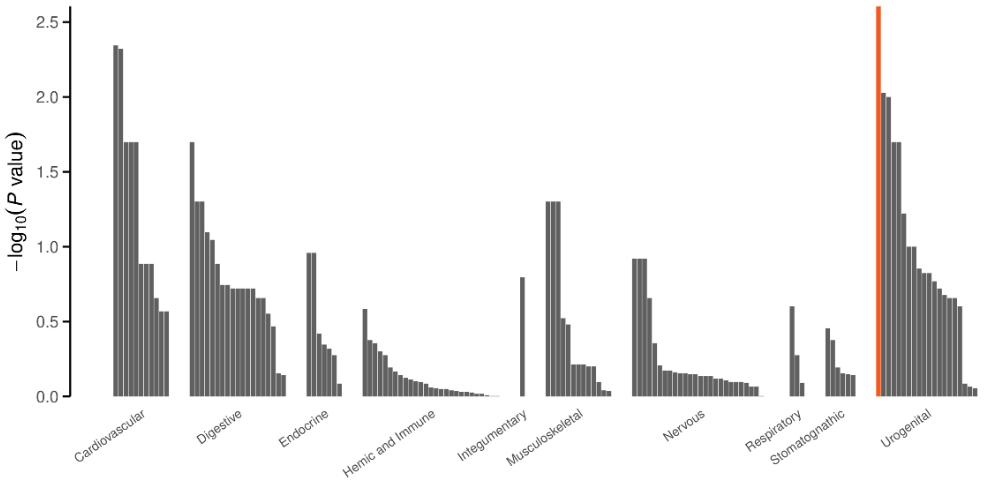


#### d. Aortic valve regurgitation fraction

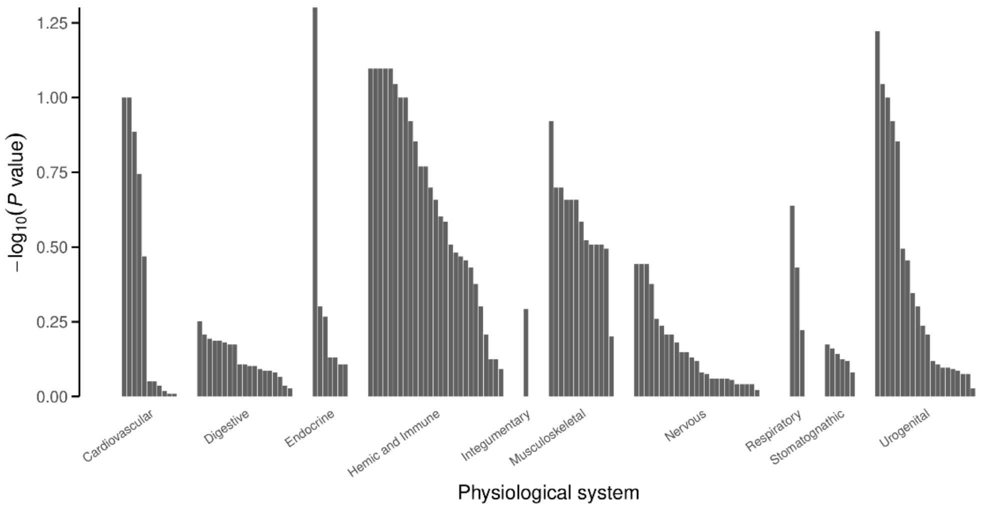

**Supplementary figure S8.** Results of the DEPICT tissue enrichment analysis for (a) Aortic annulus area (BSA indexed), (b) forward peak velocity at the aortic annulus and (c) aortic valve regurgitation fraction$_t$. The Y-axis shows the tissues clustered by the first MeSH term, ordered on the $-\log_{10}$(P-value) per cluster. The X-axis shows the $-\log_{10}$(P-value). An FDR <0.05 was considered to be statistically significant and is plotted in red and annotated, other tissues are plotted in gray.

### Supplementary discussion (SD)

#### SD1 — Epidemiological analysis and outcomes (time-to-event) analysis - complementary results

We investigated whether DeepFlow phenotyping could accurately predict clinical outcomes in aortic regurgitation in order to further validate our developed software. Complementary to the main analyses reported in our results, we also fitted a multivariate linear regression model including interaction terms to predict aortic valve regurgitation fraction considering age at cardiac MRI, sex, presence or absence of aneurysm of the aortic annulus (>= 50 mm), and presence or absence of arterial hypertension (using the I.10 code from ICD-10 codes) (Tables **S1** and **S2**). We used the R software package to perform the analysis.

**Table Supplementary discussion 1.** Results of multivariate linear regression for the prediction of aortic valve regurgitation fraction.

| Coefficients: | Estimate | Std. Error | t value | Pr(>\|t\|) | |
|---|---|---|---|---|---|
| (Intercept) | -0.085128 | 0.008173 | -10.415 | < 2e-16 | *** |
| Hypertension | 0.004184 | 0.024350 | 0.172 | 0.863566 | |
| Age | 0.099632 | 0.008197 | 12.154 | < 2e-16 | *** |
| Sex | 0.174709 | 0.012146 | 14.385 | < 2e-16 | *** |
| Aneurysm | 0.862593 | 0.172324 | 5.006 | 5.6E-07 | *** |
| Hypertension x Age | -0.048000 | 0.024581 | -1.953 | 0.050856 | . |
| Hypertension x Male | 0.006584 | 0.033047 | 0.199 | 0.842089 | |
| Age x Male | 0.023333 | 0.012079 | 1.932 | 0.053412 | . |

| | | | | | |
|---|---|---|---|---|---|
| **Hypertension x Aneurysm** | -3.392119 | 0.830656 | -4.084 | 4.44E-05 | *** |
| **Age x Aneurysm** | 0.267259 | 0.169483 | 1.577 | 0.114825 | |
| **Male x Aneurysm** | 0.496844 | 0.199183 | 2.494 | 0.012622 | * |
| **Hypertension x Age x Male** | 0.081754 | 0.032779 | 2.494 | 0.012633 | * |
| **Hypertension x Age x Aneurysm** | 3.036484 | 0.721228 | 4.210 | 2.56E-05 | *** |
| **Hypertension x Male x Aneurysm** | 2.980292 | 0.864764 | 3.446 | 0.000569 | *** |
| **Age x Male x Aneurysm** | 0.164417 | 0.198981 | 0.826 | 0.408643 | |
| **Hypertension x Age x Male x Aneurysm** | -3.060579 | 0.756937 | -4.043 | 5.28E-05 | *** |

Std. error indicates standard error; t-test: linear regression t-test; Pr(>|t|): p-value associated with the t-test. Significance codes: 0 '***' 0.001 '**' 0.01 '*' 0.05 '.' 0.1 ' ' 1. The Residual standard error was 0.9983 on 33480 degrees of freedom (6642 observations deleted due to missingness). Multiple R-squared: 0.0346, adjusted R-squared: 0.03417, F-statistic: 79.99 on 15 and 33480 DF, p-value: < 2.2e-16. A p-value < 0.05 was considered statistically significant.

**Table Supplementary discussion 2.** Residuals from the multivariate linear regression for the prediction of aortic valve regurgitation fraction.

| **Residuals** | | | | |
|---|---|---|---|---|
| **Minimum** | 1Q | Median | 3Q | Maximum |
| **-3.4214** | -0.6764 | -0.1822 | 0.4274 | 13.4159 |

1Q indicates first quartile, 3Q: third quartile.

Our multivariate linear regression shows that age at cardiac MRI, men and presence of aneurysm (but not presence of arterial hypertension) were independent predictors of higher aortic valve regurgitation fraction, which is in accordance with previous insights [14-16].

#### SD2 — GWAS sensitivity analysis

We evaluated the impact of excluding common cardiovascular comorbidities on the GWAS results of the novel traits obtained with DeepFlow (aortic valve regurgitation fraction, aortic valve regurgitation volume, net and forward left ventricular stroke volumes, peak forward and regurgitant velocities at the aortic annulus). Also, we included mitral valve regurgitation volume in the sensitivity analysis, as this trait had not yet been explored in GWAS. We followed the same quality-control steps as described in the Methods sections to perform the GWAS. Beyond the reported locus for aortic valve regurgitation fraction (LINC01808), this sensitivity analysis revealed a second independent and significantly associated genomic locus with leading SNV (7: 2,781,249 C/G), which overlaps the *GNA12* and *AMZ1* genes. Furthermore, the sensitivity analysis showed one independent leading SNV (rs3846991) significantly associated with aortic valve regurgitant volume, which also overlaps the *GNA12* and *AMZ1* genes. Moreover, the same loci could be detected for forward left ventricular stroke volume.

No new genetic loci could be found for $Ao_F$Vmax. However 16 of the reported loci for $Ao_F$Vmax and 15 of the aortic annulus area did not reach the significance level in this analysis (**Figure 1** of this supplementary discussion). This included genetic loci previously described as associated with aortic size traits in the literature (*GATA2*, *FLNB* and *SLX41P*)[12]. This decrease in significant signals is likely due to the fall in sample size (ca. 17% shrinkage) rendering inferior statistical power, combined with the complex multifactorial relationship between cardiac metrics and cardiovascular disease definitions. Mitral valve regurgitation volume and net left ventricular stroke were not significantly associated with any common genetic variant. **Figure 2** of this supplementary discussion depicts Manhattan plots for the traits with at least one significantly associated common variant.

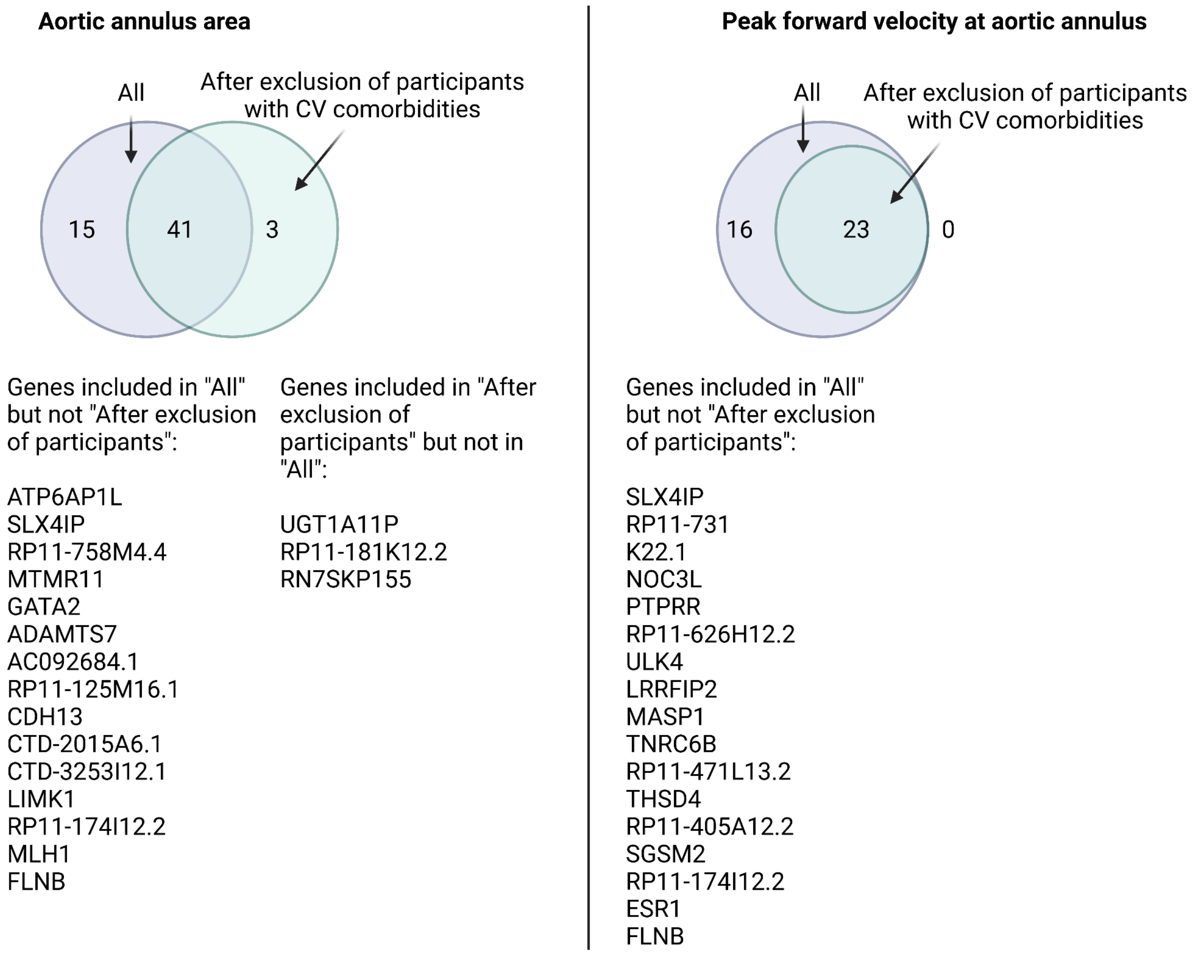


**Figure 1 Supplementary Discussion.** Venn diagrams showing the overlap of the independently associated loci after GWAS using data from all participants and GWAS after excluding participants with common cardiovascular comorbidities. The Venn diagram on the left relates to the GWAS of aortic annulus area and the Venn diagram on the right reports to the GWAS of peak forward velocity at the aortic annulus. Of note, the new loci associated with the trait of the aortic annulus area in this sensitivity analysis are non-protein encoding.

**a. Forward left ventricular stroke volume**

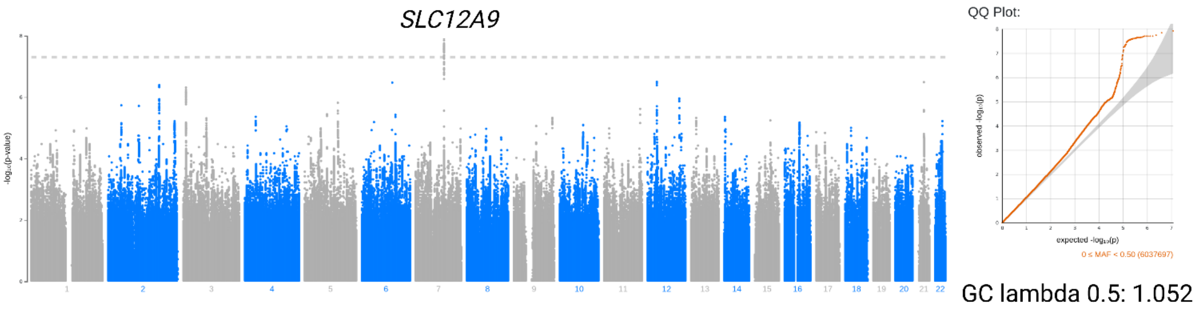


**b. Peak forward velocity at the aortic annulus**

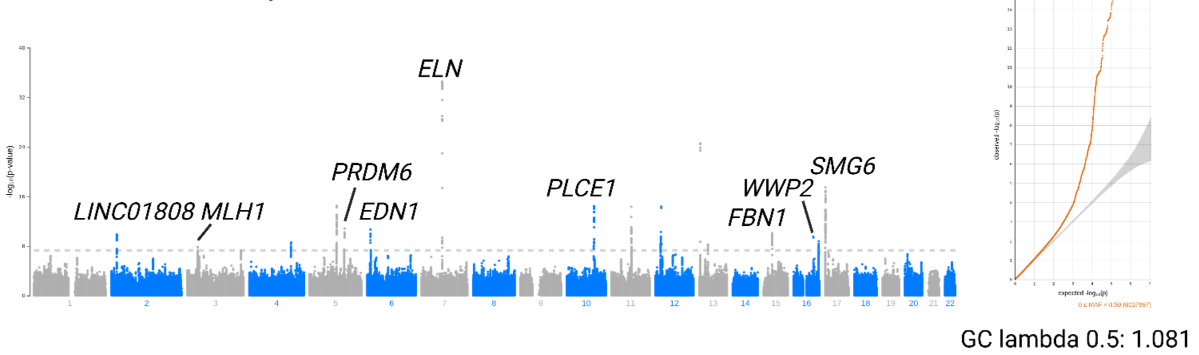


**d. Aortic annulus area**

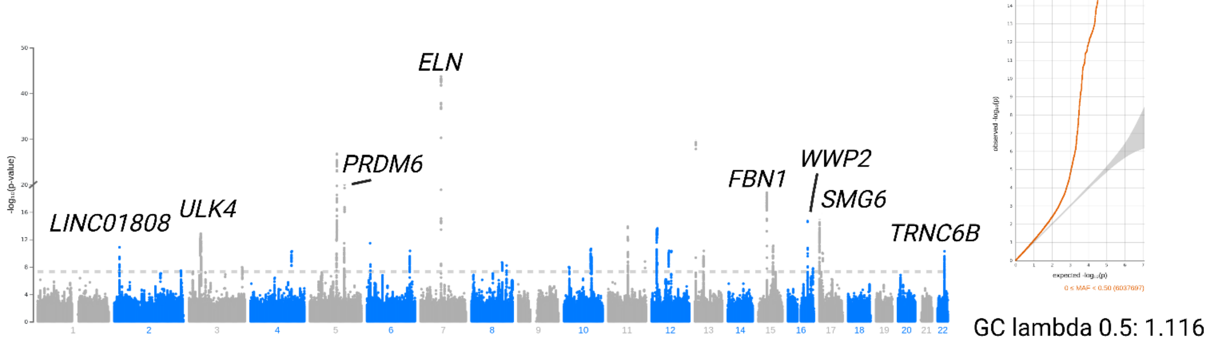


**d. Aortic valve regurgitation fraction**

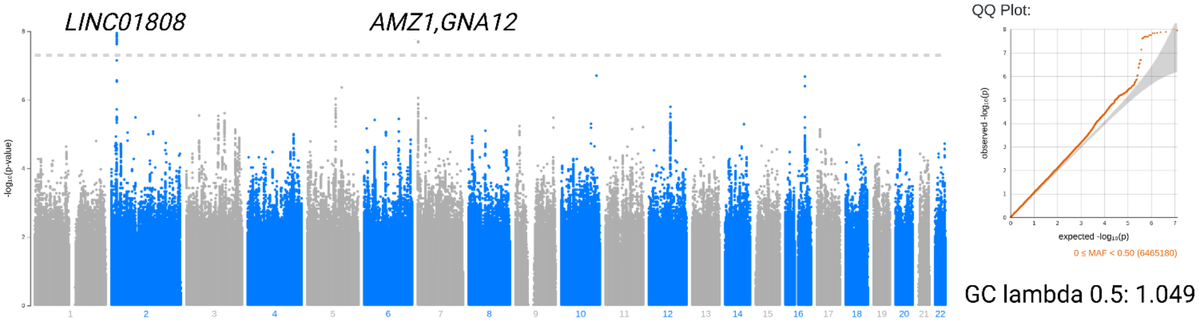

**Figure 2 Supplementary Discussion.** Manhattan plots showing the GWAS sensitivity analysis results of (A) forward left ventricular stroke volume, (B) forward peak velocity at the aortic annulus; (C) aortic annulus area; (D) aortic valve regurgitation fraction. All patients with phase contrast magnetic resonance imaging and imputed microarray DNA data. Independent variables were adjusted to age, sex and the first 5 genetic principal components. To adjust for multiple testing, we applied Bonferroni correction using the genome-wide significance threshold of $P < 5 \times 10^{-8}$ for selection of independent SNVs that were considered significantly associated with the traits. All volumes and areas are BSA-indexed. Respective QQ-plots of the phenotypes are shown. The predicted distribution in -log10 is displayed on the X-axis (P-value). The observed distribution is shown on the Y-axis as -log10 (P-value). The red dots track the observed P-values in the performed GWAS, while the gray area depicts the expected P-values from a theoretical Chi-Square-distribution. GC indicates genomic control.

#### SD3 — Flowchart regarding study sample size throughout the GWAS

We considered the first phase contrast MRI visit for this analysis. Only MRIs obeying protocol[10] (namely, with 30 frames per sequence (CINE/MAG/VENC)) were analyzed. Aortic annulus area and all volume variables were BSA indexed, so that participants with missing information regarding this indexing were excluded from the GWAS/epidemiology analysis for these traits (N of excluded participants = 2334). There were 53 phase contrast MRI with missing information regarding the time frame interval, so that net/forward left ventricular stroke volumes and aortic valve regurgitant volumes are missing in the individuals with these MRIs. Total left ventricular stroke volume is obtained through deployment of Bai et al. software. We used the same sample as for total left ventricular stroke volume on all remaining variables retrieved from Bai *et al.*'s software (N = 35633). **Figure 3** of the supplementary discussion shows a flowchart depicting the study's sample sizes throughout the GWAS study.

For the GWAS robustness/sensitivity of the traits as well as epidemiological/clinical outcomes assessment of aortic valve regurgitation fraction, participants with common cardiovascular comorbidities were excluded (see **Tables 1 to 3** for the applied ICD-10 and OPCS-4 code)**.** ICD-10 codes excluded: I.10,  I.25, I.34 to I.37, I.40 to I.43,  I.48 and I.73. Procedures excluded (using the OPCS-4 codes): K.01, K.25 to K.28, K.30, K.35, K.38, .40, K.41, K.56, K.59, K.60 and K.75.

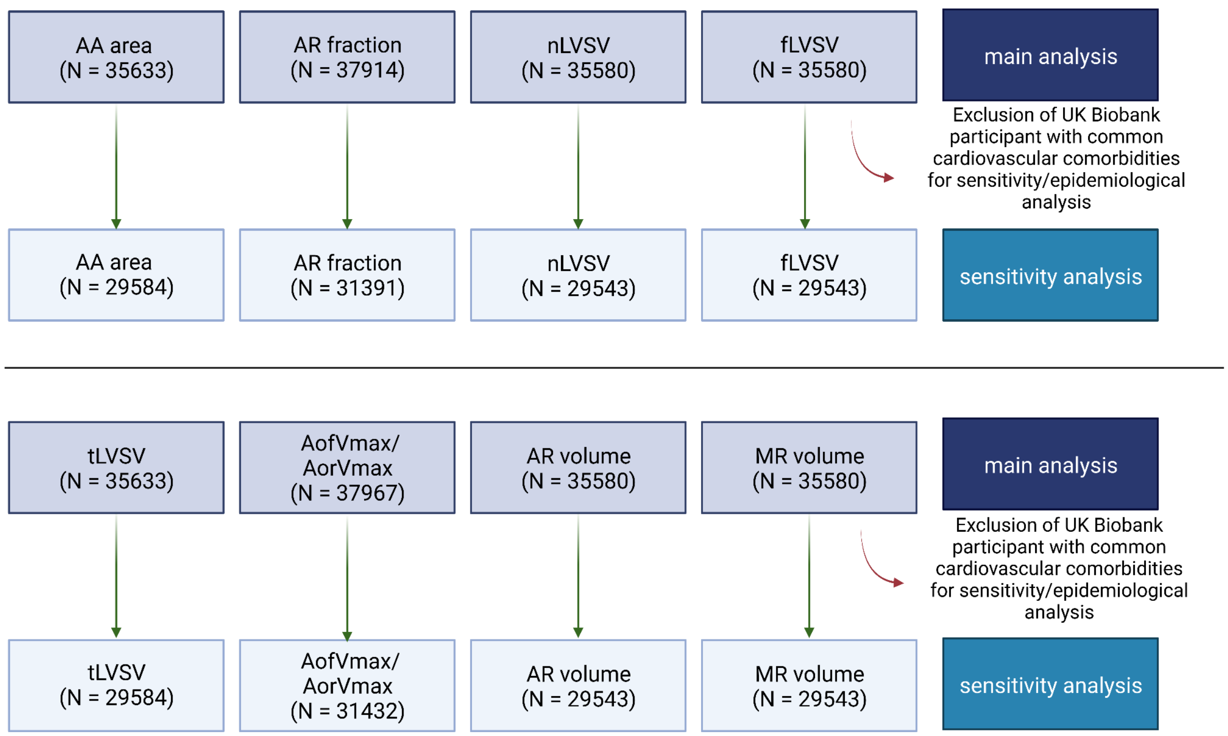


**Figure 1 Supplementary Discussion.** Flowchart regarding study sample size throughout the GWAS study. AA indicates: aortic annulus; AR: aortic valve regurgitation; fLVSV: forward left ventricular; tLVSV: total left ventricular stroke volume; nLVSV: net left ventricular stroke volume. AofVmax indicates forward peak velocity at aortic annulus. AorVmax indicates regurgitant peak velocity at aortic annulus.

### Supplementary equations

After fitting different multivariate linear regression models (see Methods) to predict aortic valve regurgitation fraction ($y$) using different combinations of covariates ($x$) (displayed in **Figure 4 of the main text**):

$a)\ y\ =\ 1.95\ +\ 0.08x,\ R^2\ =\ 0.01$. Linear regression predicting aortic valve regurgitation fraction using age at MRI as a covariate for females (**Figure 4B**).

$b)\ y\ =\ 0.76\ +\ 0.12x,\ R^2\ =\ 0.02$. Linear regression predicting aortic valve regurgitation fraction using age at MRI as a covariate for males (**Figure 4B**).

$c)\ y\ =\ -\ 14.7\ +\ 0.4x,\ R^2\ =\ 0.02$. Linear regression predicting aortic valve regurgitation fraction using age at MRI as a covariate for females with ascending aortic aneurysm (aortic annulus area >= 50 mm) (**Figure 4C**).

$d)\ y\ =\ 2.02\ +\ 0.08x,\ R^2\ =\ 0.01$. Linear regression predicting aortic valve regurgitation fraction using age at MRI as a covariate for females without ascending aortic aneurysm (aortic annulus area >= 50 mm)(**Figure 4C)**.

$e)\ y\ =\ -\ 10.9\ +\ 0.4x,\ R^2\ =\ 0.03$. Linear regression predicting aortic valve regurgitation fraction using age at MRI as a covariate for males with ascending aortic aneurysm (aortic annulus area >= 50 mm)(**Figure 4C)**.

$f)\ y\ =\ 0.93\ +\ 0.11x,\ R^2\ =\ 0.02$. Linear regression predicting aortic valve regurgitation fraction using age at MRI as a covariate for males without ascending aortic aneurysm (aortic annulus area >= 50 mm)(**Figure 4C)**.

$g)\ y\ =\ 1.73\ +\ 0.08x,\ R^2\ =\ 0.01.$ Linear regression predicting aortic valve regurgitation fraction using age at MRI as a covariate for females with hypertension (ICD-10 code I.10) (**Figure 4D)**.

$h)\ y\ =\ 4.08\ +\ 0.05x,\ R^2\ <\ 0.01.$ Linear regression predicting aortic valve regurgitation fraction using age at MRI as a covariate for females without hypertension (ICD-10 code I.10)(**Figure 4D)**.

$i)\ y\ =\ 1.55\ +\ 0.1x,\ R^2\ =\ 0.01.$ Linear regression predicting aortic valve regurgitation fraction using age at MRI as a covariate for males with hypertension (ICD-10 code I.10)(**Figure 4D)**.

$j)\ y\ =\ 0.44\ +\ 0.14x,\ R^2\ =\ 0.01.$ Linear regression predicting aortic valve regurgitation fraction using age at MRI as a covariate for males without hypertension (ICD-10 code I.10)(**Figure 4D)**.
